# Regulating brain activity in the Visual Word Form Area with real-time fMRI neurofeedback in adults with typical and poor reading skills

**DOI:** 10.64898/2025.12.08.25341820

**Authors:** N. Frei, G.S.P. Pamplona, A. Haugg, S. Brem

## Abstract

Reading proficiency is essential for everyday life, with the Visual Word Form Area (VWFA) of the brain’s reading network playing a key role in fluent reading. Reduced VWFA activation has been linked to impaired reading. Real-time functional magnetic resonance imaging (rt-fMRI) neurofeedback allows individuals to learn to regulate brain activity in targeted brain areas. While our previous study demonstrated the feasibility of VWFA regulation in adults with typical reading skills, this study investigated whether adults with poor reading skills could similarly upregulate VWFA activity using rt-fMRI neurofeedback.

Participants with typical (n = 20) and poor (n = 19) reading skills were trained to upregulate their VWFA activity during six runs of rt-fMRI neurofeedback training. Before and after training, no-feedback runs were conducted where the same regulation task had to be performed without receiving feedback. Feedback on activation was provided based on each participant’s individually defined VWFA, determined through a functional localizer task.

Our study yielded two key findings: First, upregulation of the VWFA through rt-fMRI neurofeedback can be learned irrespective of an individual’s reading proficiency. Even participants with poor reading skills, who exhibited lower VWFA activation and employed fewer reading-related mental strategies, achieved upregulation of this region through neurofeedback. Second, upregulating the VWFA through rt-fMRI neurofeedback resulted in increased VWFA activity after training, even without feedback, compared to pre-training in both adults with typical and poor reading skills.

Overall, the feasibility of regulating VWFA activity, regardless of reading skills, provides a promising foundation for developing brain-based interventions to address reading impairments through rt-fMRI-based regulation training.

## 1. Introduction

Proficient reading skills are essential for full participation in our text-driven society. However, developmental dyslexia, affecting 5-10% of the population, presents significant challenges in acquiring reading skills and performing reading tasks (Shaywitz 1998; Verhoeven 2019). Given the profound impact of developmental dyslexia on educational, career, and psychosocial development (Poskiparta et al. 2003; Mugnaini et al. 2009; Daniel et al. 2006), along with its tendency to persist into adulthood (Shaywitz et al. 1999; Shaywitz et al. 2003), novel intervention methods are urgently needed.

Our brains host a specialized cortical network dedicated to reading, known as the reading network, which comprises regions such as the inferior frontal gyrus (IFG), the intraparietal lobule (IPL), the superior temporal gyrus (STG), the left precentral gyrus (PCG), and the ventral occipitotemporal cortex (vOT) of the left hemisphere (e.g. (Martin et al. 2015)). Among these, the Visual Word Form Area (VWFA), located in the left ventral occipito-temporal cortex (lvOT), plays a pivotal role in processing written information (Dehaene and Cohen 2011; Dehaene et al. 2010; Baker et al. 2007; Glezer et al. 2009; Vinckier et al. 2007). The VWFA serves as an orthographic lexicon, housing generalized word representations that remain largely invariant to visual characteristics such as letter case and font (Wimmer et al. 2016). Its responsiveness has been linked to the perception of words and letters (Hirshorn et al. 2016; Bouhali et al. 2019), reading expertise levels (Dehaene et al. 2010; Maurer et al. 2006; Pleisch et al. 2019), and task demands (Ben-Shachar et al. 2007; Kronbichler et al. 2004). Notably, the VWFA exhibits rapid adaptation (Perrachione et al. 2016) and demonstrates activation changes in response to training effects, even within a single experimental session of letter learning through a digitized game (Pleisch et al. 2019).

Individuals who are illiterate or demonstrate poor reading skills, such as those diagnosed with developmental dyslexia, exhibit alterations in the activation and connectivity of the VWFA during reading tasks (Dehaene et al. 2010; Martin et al. 2016; Richlan et al. 2009, 2011; Maisog et al. 2008; Di Pietro et al. 2023; Brem et al. 2020; Frei et al. 2025), as well as structural differences (Linkersdörfer et al. 2012; Haugg et al. 2025) when compared to typically reading individuals. Consequently, deficient VWFA function is closely associated with poor reading fluency. The consistent alterations in VWFA function among individuals with poor reading skills (Richlan et al. 2011) underscore its significance as a potential therapeutic target for improving reading skills.

One promising approach for interventions aimed at improving dysfunctional brain function in target brain regions is functional magnetic resonance imaging (fMRI)-based neurofeedback training. This technique provides real-time feedback to participants on their brain signals in specific areas, as measured using an MRI scanner (Sitaram et al. 2017; Sulzer et al. 2013; Weiskopf 2012). This immediate feedback facilitates the learning of cognitive strategies to regulate activity or connectivity within these targeted brain regions with the aim of normalising dysfunctional brain signals to improve associated behaviour and psychiatric symptoms (Sitaram et al. 2017; Watanabe et al. 2017; Robineau et al. 2014).

In the past years, the successful use of real-time fMRI (rt-fMRI) neurofeedback to regulate functional activation has been demonstrated for numerous cortical and subcortical target brain regions (Haller et al. 2013; Berman et al. 2013; Hamilton et al. 2011; Brühl et al. 2014; Caria et al. 2007), encompassing both healthy (Habes et al. 2016; Hamilton et al. 2011; Zhu et al. 2019; Zotev et al. 2011) and psychiatric populations (Stoeckel et al. 2014; Tursic et al. 2020; Pindi). Additionally, meta-analytic studies have indicated its efficacy in ameliorating behavioural and psychiatric symptoms linked to the targeted brain areas (Dudek and Dodell-Feder 2021; Pindi et al. 2022). Moreover, and perhaps even more significantly, previous studies have demonstrated that the effects of neurofeedback training on brain activation and behaviour not only transfer to situations where feedback is unavailable but also persist over the long term. Specifically, effects have been observed to last for weeks (Subramanian et al. 2011; Yoo et al. 2008), with some cases showing continued improvement for several weeks following neurofeedback training (Rance et al. 2018).

Studies utilising rt-fMRI neurofeedback to investigate language or reading networks are notably scarce. For example, the language network, particularly the IFG and posterior STG, has been targeted using connectivity neurofeedback to alleviate auditory verbal hallucinations in patients with schizophrenia (Zweerings et al. 2019; Rota et al. 2009). These studies demonstrated that upregulation training of IFG activity resulted in enhanced language-related performance in individuals with schizophrenia. Moreover, rt-fMRI neurofeedback studies unrelated to language or reading have already been used to target specialised brain regions in the temporal lobe, such as the fusiform face area in autism spectrum disorder patients (Pereira et al. 2019) and the parahippocampal place area (Habes et al. 2016), both of which are closely situated to the VWFA. Building on these studies, our previous work (Haugg et al. 2023) demonstrated the feasibility of regulating the VWFA in the left vOT in typically reading adults using rt-fMRI neurofeedback. Furthermore, the potential of rt-fMRI neurofeedback has been explored in the context of other developmental disorders, such as Attention Deficit Hyperactivity Disorder (ADHD) (Alegria et al. 2017; Zilverstand et al. 2017) and Autism Spectrum Disorder (ASD) (Ramot et al. 2017; Direito et al. 2021).

To date, however, to the best of our knowledge, no study has used fMRI neurofeedback to target any component of the reading network in individuals with poor reading skills. These gaps emphasise the importance of further investigation into fMRI-based neurofeedback interventions for reading disorders, especially those targeting VWFA activation in dyslexia.

In our present study, we thus implemented rt-fMRI neurofeedback training targeting the VWFA in adults with poor to typical reading skills. In a first step, we aimed to replicate in our sample of adults with varying reading skills that VWFA activity, as measured during a reading task, correlates with reading proficiency, specifically pseudoword reading fluency, similar to previous research (Dehaene and Cohen 2011).

The primary aim of the study was to assess whether participants with poor reading skills could learn to upregulate activity in the VWFA through rt-fMRI neurofeedback, in a manner comparable to that of typically reading controls. We hypothesized that individuals with poor reading abilities could learn to upregulate activity in the VWFA at a comparable, or even better (see (Haugg et al. 2021)) level to individuals with typical reading skills. However, we expected individuals with poor reading skills to demonstrate reduced VWFA activation during reading tasks before neurofeedback training, in line with findings by Dehaene and colleagues (Dehaene and Cohen 2011) and Brem and colleagues (Brem et al. 2010). In addition, we aimed to explore and compare brain dynamics during self-regulation of VWFA activity with and without feedback in both individuals with typical and poor reading skills.

To assess the ability of participants with different levels of reading skill to regulate their own VWFA activity via fMRI neurofeedback, we recruited 19 adults with poor and 20 with typical reading skills. Overall, they underwent six rt-fMRI neurofeedback training runs. Each participant’s VWFA target area was identified using an individualized functional localizer. Moreover, no-feedback runs, i.e., runs without feedback, were conducted before and after the training, alongside reading fluency assessments.

Our study represents an important step in the development of novel brain-based intervention techniques for individuals with dyslexia and reading impairments. Specifically, it forms the groundwork for addressing reported hypoactivation in the VWFA during word processing (Richlan et al. 2009, 2011; Martin et al. 2016) as well as potentially enhancing reading skills through neurofeedback training. Additionally, understanding whether VWFA regulation is achievable regardless of reading skills can advance our comprehension of its causal role in reading and refine current neurobiological models.

## 2. Methods

### 2.1 Participants and Group Assignment

A total of 39 German-speaking right-handed adults (mean age 26.80 ± 5.14 years, 20 female), spanning a spectrum of reading skills from low to proficient, participated in the study. Inclusion criteria encompassed fluency in German, non-verbal IQ scores above 85 on the *Reynolds Intellectual Assessment Scales* (*RIAS NX*, (Brueggemann et al. 2006)), and the absence of psychiatric or neurological disorders. We excluded individuals displaying any magnetic resonance imaging (MRI) incompatibilities such as metal implants, pacemakers, claustrophobia, or pregnancy. All participants provided written informed consent before participation and received compensation of 60 CHF. The project received approval from the local ethics committee of the Canton of Zurich and adhered to the guidelines outlined in the Declaration of Helsinki (BASEC number 2021-00071).

Participants were divided into two groups based on their pseudoword reading fluency as measured by the *Salzburger Lese- und Rechtschreibtest* (SLRT-II, (Moll et al. 2012; Moll et al. 2014). Participants scoring above the 25th percentile were categorized as having typical reading skills (TR), whereas those scoring at or below the 25th percentile were classified as having poor reading skills (PR). This resulted in 19 participants with poor and 20 with typical reading skills. Note that the same 20 participants with typical reading skills were also included in the study conducted by Haugg and colleagues, where they were compared to participants with typical reading skills who underwent downregulation training (See (Haugg et al. 2023)).

For detailed demographic, cognitive, and reading score information, please refer to Table 1 in the Results section.

**Table 1.** Descriptive and demographic measures for participants with typical and poor reading skills.

|  | TR | PR | PR vs. TR |  |
| --- | --- | --- | --- | --- |
| | | | t-test t(DoF) or chi-squared test $\chi^2$ (DoF) | |
| | Mean ± SD | | t(DoF)/ $\chi^2$ (DoF) | p-values |
| **Demography** |  |  |  |  |
| Participants | 20 | 19 |  |  |
| Sex (female/male) | 11/9 | 9/10 | $\chi^2(1) = 0.23$ | 0.634 |
| Handedness (EHI) | 78.18 ± 15.1 [45.45, 100] | 77.63 ± 24.48 [0, 100] | $t(36) = 0.081$ | 0.936 |
| Age (in years) | 26.76 ± 5.01 [18.17, 43.5] | 26.85 ± 5.13 [19.92, 42.33] | $t(37) = -0.50$ | 0.960 |
| Years of schooling | 15.97 ± 4.05 [11, 20] | 16.68 ± 3.5 [12, 27] | $t(37) = -0.57$ | 0.573 |
| ARHQ (mean) | 8.64 ± 2.16 [0.5, 10.91] | 9.44 ± 2.34 [3.08, 12.9] | $t(37) = -1.08$ | 0.286 |
| Non-verbal IQ (RIAS-NX) | 110.45 ± 5.58 [100, 121] | 102.53 ± 15.65 [88, 117] | $t(37) = 2.07$ | 0.045 |
| **Reading-related skills** |  |  |  |  |
| Reading fluency words (SLRT-II perc) | 69.03 ± 19.35 [30, 90] | 16 ± 13.39 [1.5, 47.5] | $t(37) = 9.65$ | <0.001* |
| Reading fluency pseudowords (SLRT-II perc) | 73.5 ± 17.23 [48, 97.5] | 15.21 ± 6.43 [0, 25] | $t(24.44) = 13.77$ | <0.001* |
| Mean words&pseudowords reading fluency (SLRT-II perc mean) | 71.26 ± 16.43 [39, 98.25] | 15.63 ± 8.79 [0.75, 32.75] | $t(29.38) = 12.93$ | <0.001* |
| **Neurofeedback-related measures** |  |  |  |  |
| Motivation before training (on 1-10) | 8.65 ± 1.19 [5, 10] | 8.32 ± 1.38 [5, 10] | $t(37) = 0.79$ | 0.435 |
| Tiredness before training (on 1-10) | 3.7 ± 1.79 [1, 7] | 3.95 ± 1.61 [2, 7] | $t(37) = -0.44$ | 0.661 |
| Attention during training (on 1-20, mean) | 13.58 ± 3.69 [2, 17.33] | 13.41 ± 2.48 [6.83, 17.5] | $t(37) = 0.16$ | 0.873 |
| Number of different strategies used | 2.05 ± 0.80 [1, 4] | 2.05 ± 1 [1, 4] | $t(37) = -0.01$ | 0.993 |
| Proportion of “imagine reading” as a strategy in NF runs (%) | 100 | 65 | $\chi^2(1) = 10.37$ | 0.005* |
| Proportion of “imagine reading” as a strategy in post-training runs (%) | 17 | 13 | $\chi^2(1) = 1.51$ | 0.273 |

### 2.2 Experimental Design

#### 2.2.1 Procedure

Before the experiment, participants underwent a brief screening and cognitive assessments, including reading-related tests. Following this, they participated in a 1-hour MRI session encompassing both structural and functional measurements. During this session, participants underwent a functional localizer task designed to identify the VWFA on an individual basis. Additionally, they performed a neural adaptation task (c.f. (Perrachione et al. 2016)), which was not analysed in this study. Next, participants engaged in a real-time functional MRI (fMRI) neurofeedback training session, comprised of six runs. Before neurofeedback training, participants completed a no-feedback run, during which they performed the same task as in the feedback runs but received no performance-related feedback. After neurofeedback training, participants repeated the no-feedback run, the functional localizer, and the neural adaptation task. Finally, participants again underwent reading-related tests outside the scanner (Figure 1).

**Figure 1.**
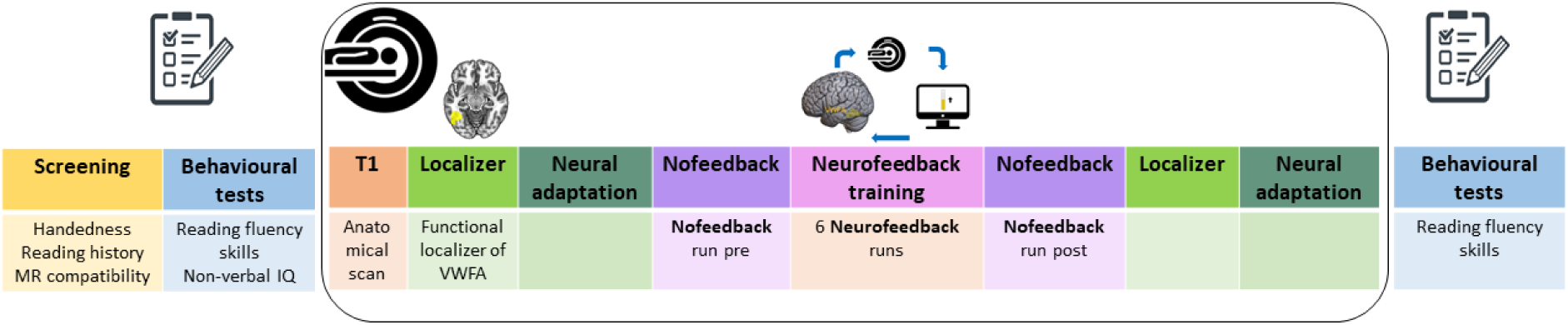
Experimental design. After a brief screening, participants underwent behavioural tests to assess reading fluency and non-verbal intelligence. During the MRI session, participants underwent a T1-weighted anatomical scan, a localizer, a neural adaptation run, a no-feedback run, and neurofeedback training runs. After neurofeedback training, participants repeated the no-feedback, neural adaptation, and functional localizer runs inside the MRI scanner, as well as the reading fluency test outside the scanner.

#### 2.2.2 Screening and Cognitive Assessments

Behavioural screening and administered tests were conducted via phone, at home, and an online web application (REDCap, https://www.project-redcap.org/). This screening encompassed an assessment of handedness (*Edinburgh Handedness Inventory* (EHI; (Oldfield 1971)), completion of the *Adult Reading History Questionnaire* (ARHQ; (Lefly and Pennington 2000)), and a collection of basic demographic information. The cognitive assessment involved measurements of non-verbal IQ using two subtests from the German version of the *RIAS NX* (i.e., the “*Odd-item out”* and “*What is missing?”* subtests). Reading fluency was assessed using the SLRT-II, which quantified performance by counting the number of words or pseudowords correctly read within one minute. As our study involved adult participants, only pseudoword reading fluency was used for analyses involving reading fluency scores. Pseudowords are artificial combinations of letters that adhere to the phonotactic rules of a language but do not represent meaningful words. A focus on pseudowords provides more discernible insights into reading impairments among adults, as pseudowords, unlike real words, which may allow for compensatory strategies developed over time, reveal difficulties in the automatic mapping of letters to corresponding speech sounds. Finally, participants rated their motivation to participate and their level of tiredness on a scale ranging from one to ten both at the beginning and at the end of the MRI session.

#### 2.2.3 fMRI Tasks

During MRI recordings, multiple tasks were performed. Following an anatomical scan, several functional tasks were conducted, including no-feedback and neurofeedback runs, and a localizer task to identify the VWFA (details provided below). All functional tasks were repeated after the neurofeedback training.

##### 2.2.3.1. Functional Localizer of the VWFA

The localizer task was used to determine the location of the VWFA for each participant. This individually defined region was then used as the target region for neurofeedback training. The localizer task consisted of nine blocks of word presentations interleaved with nine blocks of checkerboard presentations. A fixation cross was presented for 2 seconds between all blocks. Each word block lasted 16.5 seconds, during which 15 German words were displayed for 800 milliseconds each, with an inter-stimulus interval of 300 milliseconds (Figure 2). The presented words were all nouns and selected based on the number of graphemes (5–8), syllables (2–3), and word frequency (ranging from seven to 10,000 words per million), as assessed by the WordGen software ((Duyck et al. 2004), https://www.wouterduyck.be/wordgen, which uses the CELEX and Lexique lexical databases for word selection. Checkerboards were presented for the same duration as the words. During the localizer task, participants were instructed to press a button on a response pad (Cambridge Research Systems, UK) whenever either a word or bars above and below the checkerboard were displayed in red. The button press task was performed to assess compliance and attention.

**Figure 2.**
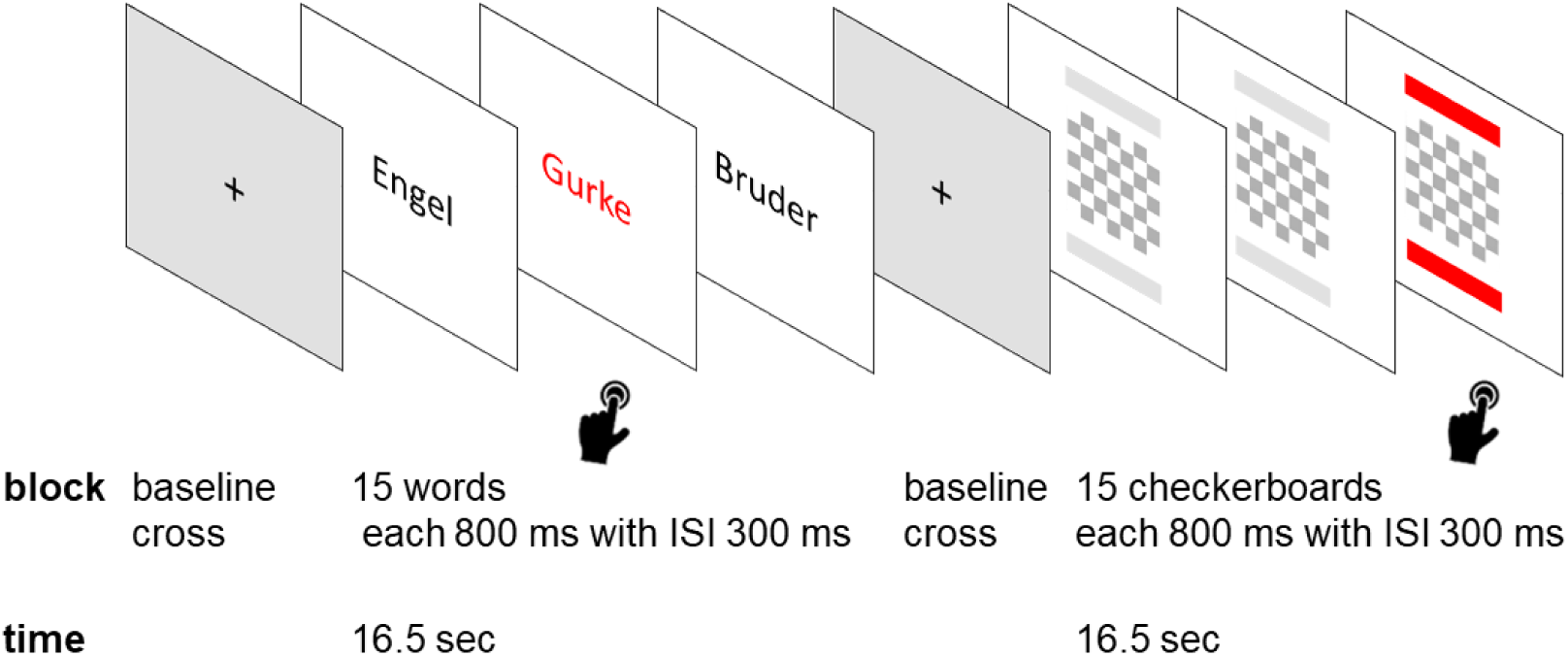
Functional localizer paradigm to identify the location of the Visual Word Form Area. The task consisted of nine blocks of word presentations alternated with nine blocks of checkerboard presentations. Additionally, a 2-second baseline cross was presented between the blocks. Each word block lasted 16.5 seconds, during which 15 German nouns were sequentially displayed for 800 milliseconds each, separated by inter-stimulus intervals (ISI) of 300 milliseconds. During the task, participants were instructed to press a button whenever a word or the bars under and above the checkerboard were displayed in red.

To create a mask of each participant’s VWFA, we selected the 50% most active voxels during the localizer task within a predefined VWFA mask. This predefined mask was constructed based on coordinates from eight different studies (see supplementary material Chapter 1.1 with Table S1 and Figure S1 for a detailed description; (Haugg et al. 2023)). These coordinates were selected to encompass a large portion of the area associated with the VWFA (the used mask can be accessed via our Open Science Framework page: https://osf.io/w3xzv/).

##### 2.2.3.2. Neurofeedback and No-feedback Paradigms

The rt-fMRI neurofeedback training comprised six runs, each with a duration of 4 minutes. Within each run, participants engaged in four regulation blocks of 40 seconds, interspersed with four baseline blocks of 20 seconds. During the regulation blocks, participants were instructed to upregulate their VWFA activity, represented by the level and colour of a thermometer icon, using a reading-related mental strategy. Participants were explicitly encouraged to identify and evaluate successful reading-related strategies throughout the experiment. They were asked to adjust or change their reading-related mental strategy for the next neurofeedback run based on their displayed feedback and to select the most effective strategy for the final no-feedback run. Feedback was continuously presented to the participants and displayed via goggles. Feedback was presented using Psychopy 2.7 https://www.psychopy.org/. Participants were briefed on the VWFA’s role within the reading network and the inherent latency of 5-7 seconds in fMRI neurofeedback. During baseline blocks, participants were instructed to engage in imagery of playing tennis (Figure 3) as a way to interrupt cognitive engagement with reading-related strategies. Before and after each neurofeedback run, participants were asked about the strategies they planned and used to up-regulate their VWFA activity. Participants rated their perceived level of attention and performance on a scale ranging from 1 (very low) to 20 (very high) after each neuro- and no-feedback run.

**Figure 3.**
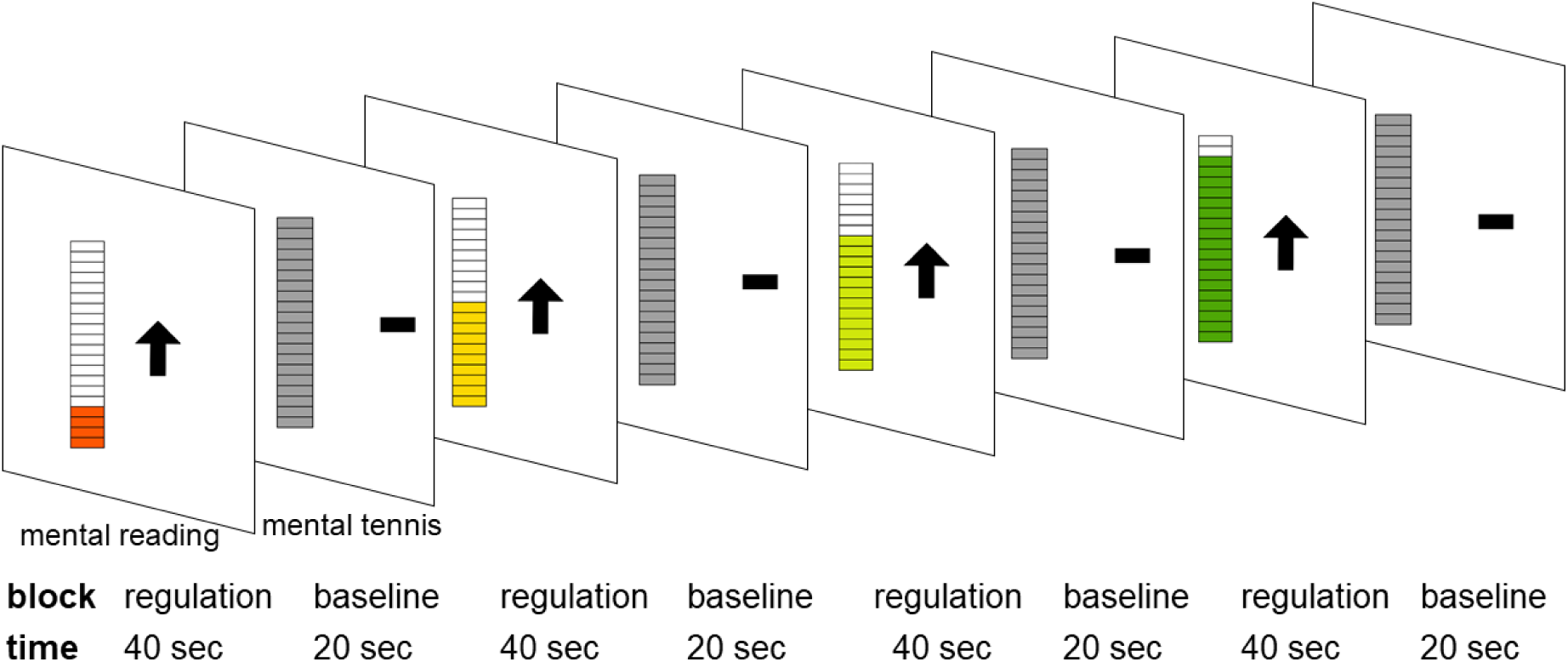
**Neurofeedback paradigm**. During the four regulation blocks, participants were instructed to engage in reading-related mental strategies aimed at upregulating activity in their visual word form area. Conversely, during the four baseline blocks interspersed with the regulation blocks, participants were instructed to mentally engage in playing tennis to divert their attention away from reading-related strategies.

The structure of the no-feedback runs mirrored that of the neurofeedback runs. However, participants did not receive any feedback on their VWFA activity during regulation blocks, allowing for an exploration of their performance in the absence of feedback. Thus, stable blue thermometer levels were displayed during the regulation blocks (please refer to supplementary material, Figure S2, for the no-feedback paradigm).

#### 2.2.4 MRI Acquisition

MRI data were recorded at the University Hospital of Psychiatry in Zurich using a Philips Achieva 3 Tesla scanner (Philips Medical System, Best, The Netherlands) equipped with a 32-channel head coil. Functional images for the localizer and neural adaptation runs were acquired using a T2*-weighted, multiband echo-planar imaging (EPI) pulse sequence comprising 247 volumes covering the whole brain. The sequence parameters were: repetition time (TR) = 1.25 s, echo time (TE) = 35 ms, flip angle = 80°, field of view (FOV) = 197 x 197 x 138 mm^3^, isotropic voxel size = 3 mm^3^, slice gap = 0.3 mm, 42 ascending slices, and a multi-band factor = 2. The neuro- and no-feedback runs comprised of T2*-weighted images totaling 135 volumes, acquired with the following sequence parameters: TR = 2 s, TE = 35 ms, flip angle = 82°, FOV = 220 × 220 × 110 mm³, voxel size = 2 × 2 x 3 mm^3^, slice gap = 1 mm, 27 ascending slices. In all functional runs, five dummy scans were acquired before the functional acquisition to allow for steady-state magnetisation.

Additionally, high-resolution T1-weighted anatomical images were acquired for each participant using a magnetisation-prepared rapid acquisition gradient echo (MPRAGE) sequence with the following parameters: TR = 6.8 s, TE = 3.2 s, flip angle = 9°, FOV = 270×255×176 mm^3^, isotropic voxel size of 1 mm³, and 176 slices.

### 2.3 Data analysis

#### 2.3.1 Statistical Analysis of Behavioural Measures

Two-sample *t*-tests were performed to compare behavioural and demographic measures between participants with poor and typical reading skills (Table 1). Additionally, pseudoword reading fluency tests conducted before and after neurofeedback training for both groups were analysed using a mixed ANOVA with timepoint (pre/post) as within-subject factor and group (TR, PR) as between-subject factor (supplementary material Chapter 2.5, Figure S8). Behavioural analyses were performed with IBM SPSS 24 https://www.ibm.com/de-de/products/spss-statistics.

#### 2.3.2 Statistical Analysis of fMRI Data

##### 2.3.2.1. Preprocessing

MRI data underwent preprocessing using the Statistical Parametric Mapping (SPM12) toolbox, developed by the Wellcome Trust Centre for Neuroimaging at University College London, UK (http://www.fil.ion.ucl.ac.uk/spm), running on MATLAB (version R2022b; https://www.mathworks.com/products/matlab.html).

As neurofeedback training was performed in native space rather than in Montreal Neurological Institute (MNI) space, the functional localizer, neurofeedback runs, and no-feedback runs were analysed both in native space and in MNI space. Data in native space enabled the investigation of signals within the exact neurofeedback target ROI, while data in MNI space allowed for group-level analyses.

The preprocessing pipeline in MNI space included slice-time correction, realignment, coregistration, segmentation, normalisation to MNI space, and smoothing with a 6-mm full width at half maximum kernel. The preprocessing pipeline in native space included slice-time correction, realignment, coregistration of the anatomy to the mean functional image, and smoothing with a 6-mm full width at half maximum kernel.

##### 2.3.2.2. Functional Localizer of the VWFA Localisation of each participant’s VWFA

Functional localisation of the VWFA in the left occipito-temporal cortex was performed on a single-subject level and in each participant’s non-normalised native space. After the preprocessing in native space, a general linear model (GLM) with regressors for words, checkerboards, and the six motion parameters was calculated. The neurofeedback target mask was then calculated as the 50% most active voxels of the words_vs_checkerboard contrast within a larger VWFA mask created based on literature (Table S1 and Figure S1).

###### Correlation between VWFA activity and reading fluency

To assess the association between VWFA activity and reading skills, we calculated the Pearson correlation coefficient between pseudoword reading fluency scores (expressed as percentiles) and VWFA activity. VWFA activity was defined as the mean contrast value for the words_vs_checkerboard contrast of a participant’s non-normalised native space within the VWFA. To replicate this analysis in a larger group, we also calculated the same Pearson correlation using 18 additional participants with typical reading skills from our previous study (Haugg et al. 2023) (Supplementary material Figure S3).

###### Whole-brain group-level activity during the functional localizer run

Finally, to assess group-level activity, we performed additional analyses with data preprocessed in MNI space. On the first level, we again calculated a GLM with regressors for words, checkerboards, and the six motion parameters. Then, we performed three second-level GLMs for typical readers, poor readers, and the contrast between typical and poor readers (Supplementary material Figure S4, Tables S4, S5). A cluster-defining threshold of *p*_CDT_ = 0.001 and cluster-level threshold of *p*_FWEc_ < 0.05, family-wise error corrected, were employed across all three analyses.

##### 2.3.2.3. Neurofeedback training and no-feedback Runs

###### Neurofeedback online analysis

To compute feedback signals during neurofeedback training, fMRI data were analysed in real-time. This analysis was conducted using the OpenNFT toolbox (http://opennft.org/) (Koush et al. 2017b; Koush et al. 2017a). OpenNFT received the functional volumes in real-time via the DRIN export system. All fMRI data underwent preprocessing using OpenNFT’s default pipeline, which includes real-time realignment to account for motion and spatial smoothing with a Gaussian kernel of 5mm full width at half maximum. Additionally, default denoising was applied, which included drift removal using a cumulative GLM, Kalman filtering for temporal smoothing, and a first-order autoregressive model to remove temporal correlations. Subsequently, OpenNFT’s default dynamic scaling approach ensured that participants did not experience ceiling or floor effects in activity regulation. The corrected signal was converted to a value between 1 (low signal) and 20 (high signal) and subsequently provided to the participant as real-time feedback. Feedback was presented in the form of a thermometer that changed colour from red to yellow to green, visually indicating the level increase corresponding to the level of activation in 20 increments.

###### Offline fMRI analyses of neuro- and no-feedback runs

For offline analyses after neurofeedback training, we performed standard preprocessing, including normalisation (see 2.3.2.1). Then, we performed first-level analyses using a GLM that included regressors for regulation, baseline (playing tennis), and the six motion regressors. Additionally, motion censoring was implemented to address excessive motion, identified by framewise displacement values exceeding 0.9 mm in individual images. Then, over all neurofeedback runs, second-level GLMs were conducted. Statistical significance was assessed using a two-sample t-test to investigate differences between the two groups during neurofeedback training runs, alongside one-sample t-tests conducted for each group separately. A cluster-defining threshold of *p*_CDT_ = 0.001 and cluster-level threshold of *p*_FWEc_ < 0.05, family-wise error corrected, were employed across all three analyses.

The same analyses as the neurofeedback offline analyses were conducted for the two no-feedback runs. Here, second-level GLMs were conducted to analyse pre- and post-training runs. One-sample t-tests were used for each group separately, two-sample t-tests for between-group comparisons, and paired t-tests to assess within-group changes from pre- to post-training.

###### Percent signal change analyses

Finally, to investigate dynamic signal progressions within the VWFA, we extracted percent signal change (PSC) time courses during neurofeedback and no-feedback runs using MarsBaR (http://marsbar.sourceforge.net). For this extraction, we utilised a VWFA mask based on literature, as previously described (Haugg et al. 2023) (Figure S1 and Table S1).

Following the neurofeedback analyses by MacInnes and colleagues (MacInnes et al. 2016), we investigated three different factors: group (typical readers, poor readers) as a between-subject factor, phase (early: first half of a regulation block, late: second half of a regulation block) as a within-subject factor, and timepoint (before neurofeedback, during neurofeedback, after neurofeedback) as within-subject factor. The three factors were analysed using a mixed ANOVA. Splitting the time courses into an early phase (first 20 seconds/10 TRs (repetition time) and a late phase (last 20 seconds/10 TRs (repetition time)) allowed us to investigate whether self-regulation with and without feedback could be sustained over longer periods. Post-hoc analysis used the emmeans library in R. We adjusted *p*-values in post-hoc tests using the pairwise comparisons correction (PWC2; (Shaffer 1995)) method.

Parallel to MacInnes et al. (MacInnes et al. 2016), we also investigated whether VWFA activation during different phases and timepoints was significantly different from baseline. We investigated this research question using one-sample t-tests against zero for each participant group. In addition, we performed two-sample t-tests to compare VWFA activity in participants with poor and typical reading skills during each phase and at each timepoint.

Finally, to investigate the specificity of our effects, we repeated our analyses with a control region unrelated to reading, the foot area of the motor cortex (MC foot). To define this region, we used Neurosynth (Yarkoni et al., 2011; https://neurosynth.org/) using the term “foot”. The peak of the cluster associated with “foot” was located in the motor cortex (−2, −30, 74). In addition to the MC foot, we also investigated activation during neuro- and no-feedback runs in other areas of the reading network, and we repeated our analyses for additional key regions associated with reading. Those additional regions of the reading network in the left hemisphere were selected based on prior studies. These included the superior temporal gyrus (STG, MNI: −60, −32, 6 mm), the inferior frontal gyrus (IFG, −56, 12, 15), the precentral gyrus (PreG, −44, 6, 30), and the inferior parietal lobule (IPL, −40, −48, 42) (Cao et al., 2008; Morken et al., 2017; Wise Younger et al., 2017). The ROIs were defined as spheres of 8 mm (for the regions of the reading network) and 6 mm (for the MC foot) around the peak coordinates using MarsBaR.

Corrections for multiple comparisons were applied using the False Discovery Rate (FDR) method (Benjamini and Hochberg 1995).

For additional exploratory analyses of the activation progression during the phase of sustained activity (late phase), please refer to the supplementary material, Chapter 2.3. All statistical analyses were performed with IBM SPSS 24 and RStudio (R version 4.4.0; https://www.rstudio.com/).

##### 2.3.2.4 Evaluation of the Mental Strategies

To analyse the individual strategies employed during neurofeedback training, we categorized the strategies into six subgroups (Table S2, Figure S7). This classification was conducted independently by eleven raters, each tasked with sorting the reported strategies into six predefined categories of imagination strategies: *reading text*, *reading words*, *reading letters*, *writing*, *thinking in spoken language*, and *others*. The final classification for each strategy was determined by majority consensus among the raters. For further analyses, reading-related strategies (‘reading text’, ‘reading words’, ‘reading letters’) were grouped as “imagine reading”, while less reading-related strategies (writing, thinking in spoken language, others) were categorised as “imaging other”. A chi-square test was performed to determine whether participants with typical reading skills employed “imagine reading” strategies more frequently than those with poor reading skills during the six neurofeedback runs and in the post no-feedback run.

## 3. Results

### 3.1 Behavioural and Demographic Measures

Descriptive statistics for behavioural and demographic measures in participants with typical and poor reading skills, along with comparisons between these groups, are shown in Table 1.

Importantly, no significant differences were observed between the two groups in measures that could influence the success of neurofeedback training. Specifically, self-rated tiredness (*t*(37) = −0.44, *p* = 0.661) or self-rated motivation prior to the training (*t*(37) = 0.79, *p* = 0.435) did not differ significantly. Similarly, self-rated attention scores during the training sessions were comparable between the groups (*t*(37) = 0.16, *p* = 0.873).

As per definition, the group of participants with poor reading skills showed significantly lower scores in reading fluency than the group of typical readers (*t*(29.38) = 12.93, *p* < 0.001).

Non-verbal intelligence (Reynolds Intellectual Assessment Scales (RIAS_NX percentage)), handedness (Edinburgh Handedness Inventory (EHI) and reading fluency (Salzburger Lese- und Rechtschreibtest (SLRT-II)) are standardized measures. ARHQ (Adult Reading History Questionnaire) is a reading history inventory. Motivation and tiredness were self-rated by the participants on a scale of 1-10 before training and attention was self-rated on a scale of 1-20 after each neurofeedback run. Descriptive statistics (*n* = 39) are mean ± standard deviation (SD). Group comparisons (TR vs. PR) are *t*-values and chi-squared values (DoF = degree of freedom) and *p*-values. * significant values (p < 0.05) after correction for multiple comparisons with FDR. perc = percentile, [] = range.

### 3.2 Functional localizer run

#### 3.2.1. VWFA localization

The functional localizer run allowed us to localize the individual VWFA in all participants. Specifically, all participants showed positive mean values within the VWFA for the words_vs_checkboards contrast. In addition, each of the 20 participants with typical reading skills showed significant activation (whole-brain initial cluster defining threshold *pCDT* = 0.001, cluster-level correction at *pFWEc* < 0.05) in the VWFA when performing whole-brain analyses. In poor readers, VWFA activity was lower, and most participants did not show significant activation in the VWFA when performing whole-brain analyses. However, when focusing these analyses on the VWFA with small volume correction, all but one participant with poor reading skills showed significant activation (whole-brain initial cluster defining threshold *pCDT* = 0.001, small volume correction at *pFWEc* < 0.05) (Figure 4 and Table S3). Notably, neurofeedback training remains feasible even with non-significant but positive activation observed in this region.

**Figure 4.**
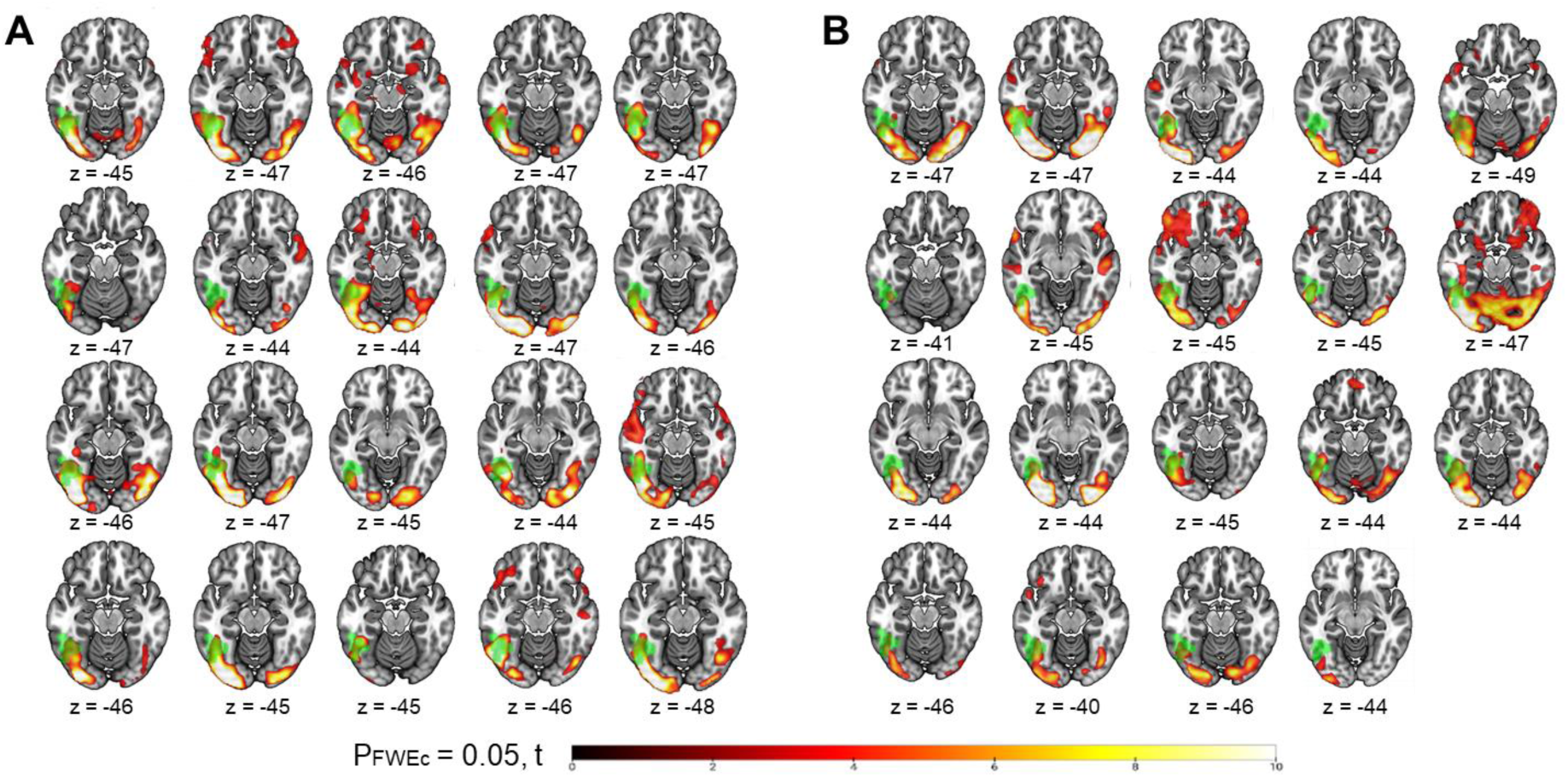
Whole-brain activation for each participant in **(A)** adults with typical reading skills and **(B)** adults with poor reading skills during the functional VWFA localizer run. Initial cluster defining threshold *p_CDT_* = 0.001, *p*_FWEc_ < 0.05, significant activation in hot colours, VWFA mask in green. The participant with no significant activation in the VWFA is in the lowest row on the right.

On a group level, we found that both groups, i.e. participants with typical and participants with poor reading skills, exhibited significant positive activity in the VWFA during the functional localizer run prior to neurofeedback training (Figure S4 and Tables S4 and S5). Additionally, both groups also showed activity in the bilateral inferior occipital gyrus and right precentral gyrus.

#### 3.2.2. Correlation between VWFA activity and reading fluency

We found a significant positive correlation between pseudoword reading fluency (SLRT-II percentile) and VWFA activity (mean value of the words_vs_checkerboards contrast within the VWFA) during the functional localizer run (r = 0.383, p = 0.016, n = 39) (Figure 5). To further verify this finding, we repeated this analysis using a larger sample, which included data from 18 additional adults who completed the same task (described in Haugg et al. (2023)). This additional analysis in an enlarged sample confirmed the positive correlation between pseudoword reading fluency and VWFA activation (r = 0.523, p < 0.001, n = 57) (Figure S3).

**Figure 5.**
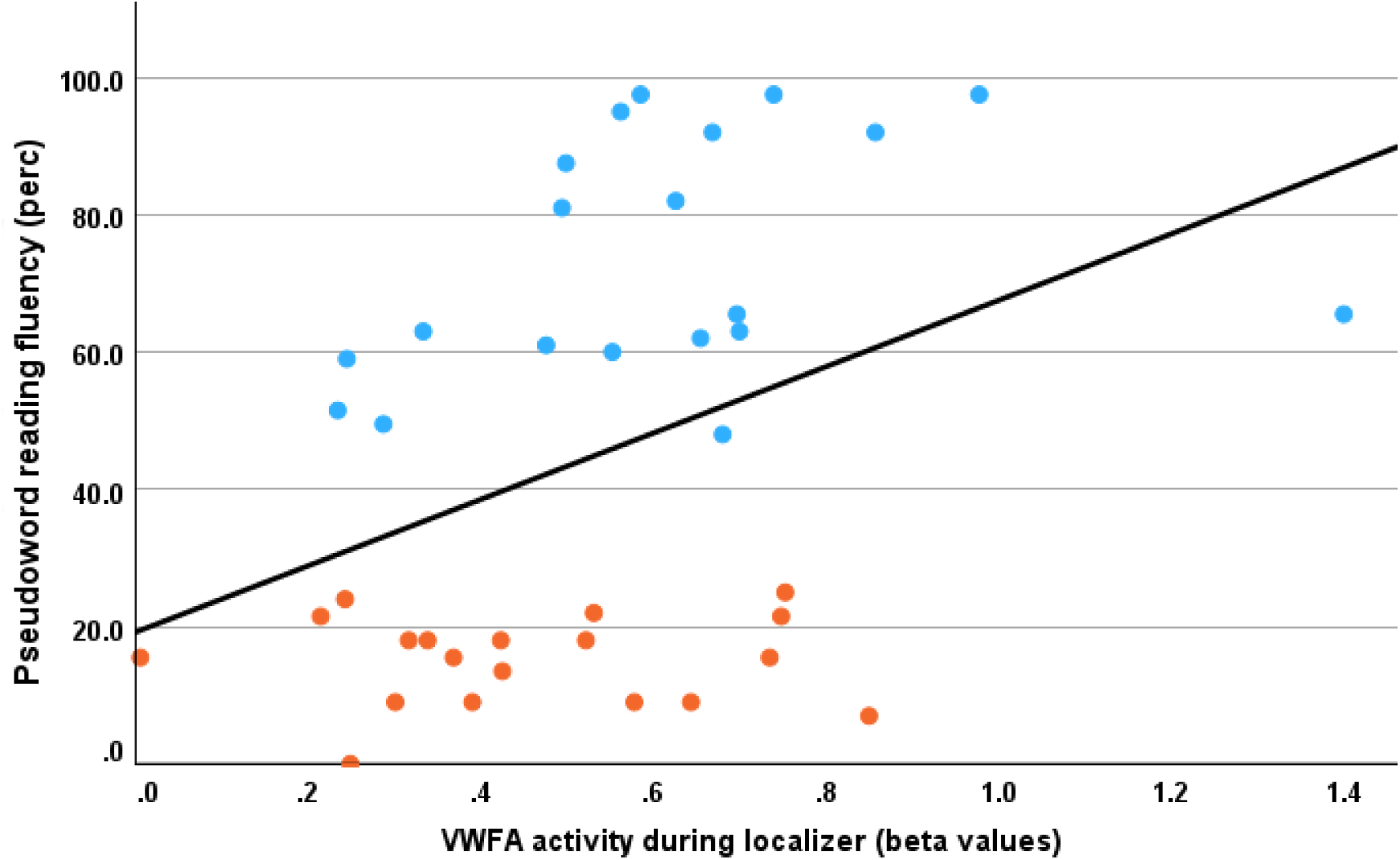
A significant positive correlation was observed (r = 0.38, *p* = 0.016, *n* = 39) between the percentile (perc) score of pseudoword reading fluency (SLRT-II) and the beta values of the VWFA for the contrast words vs checkerboards. Participants with typical reading skills are shown in blue, while those with poor reading skills are shown in red.

### 3.3 Brain Activation during Neurofeedback Training Runs

A second-level analysis across all six neurofeedback training runs revealed significant up-regulation of the VWFA in participants with both typical and poor reading skills (Figure 6). In addition, we found significant activation in other regions of the reading network, including the left STG, MTG, precentral gyrus (PrecG), angular gyrus (AnG), and left IFG during the regulation phase compared to baseline (Figure 6 and Tables 2 and 3). Further significant activation clusters were found in the striatum (Str), anterior insula (AI), sensorimotor cortex (SMC), inferior occipital gyrus (IOG), and middle frontal gyrus (MFG). There were no significant group differences between participants with typical and poor reading skills in the whole-brain analysis during neurofeedback.

**Figure 6.**
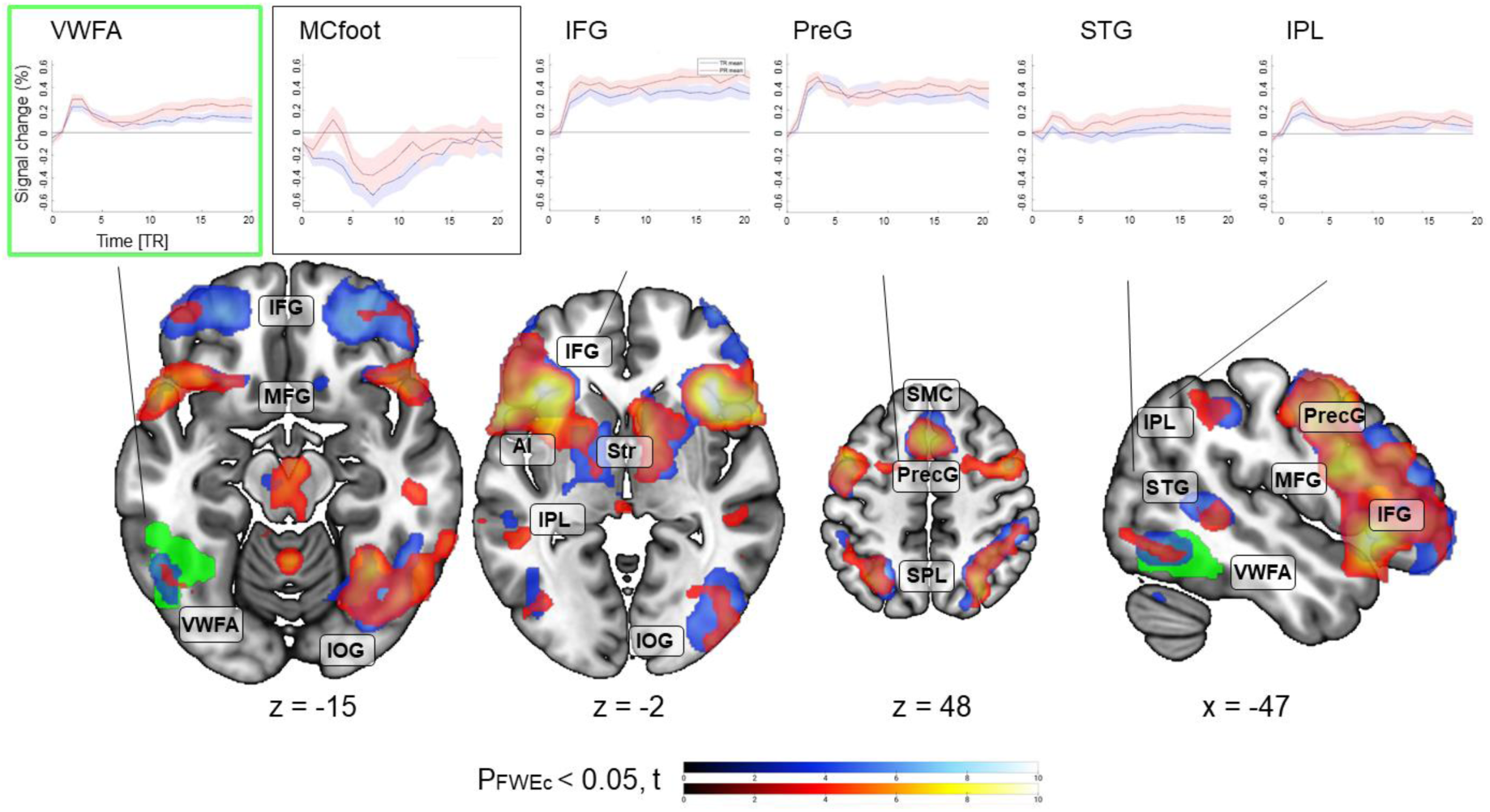
Activations for the contrast regulation vs baseline across all six neurofeedback runs for typical (cold colours) and poor (warm colours) readers. Significant activations were observed in bilateral anterior insulae (AI), bilateral inferior frontal gyri (IFG), middle temporal gyrus (MTG), left superior parietal lobe (SPL), angular gyrus (AnG), left inferior occipital gyrus (IOG), visual word form area (VWFA), right middle temporal gyrus (MTG), left supramarginal cortex (SMC), bilateral precentral gyri (PrecG), striatum (Str). Green: VWFA mask. Cluster defining threshold *p_CDT_* = 0.001, *pFWEc* < 0.05. On top, the PSC time courses during the neurofeedback regulation blocks are shown (mean over 6 runs) for typical (blue) and poor (red) readers. PSC time courses during no-feedback runs are shown in supplementary material Figure S6. For further details on the PSC time courses of the VWFA and MC foot during neurofeedback training, see Figure 7A and B.

**Table 2.**
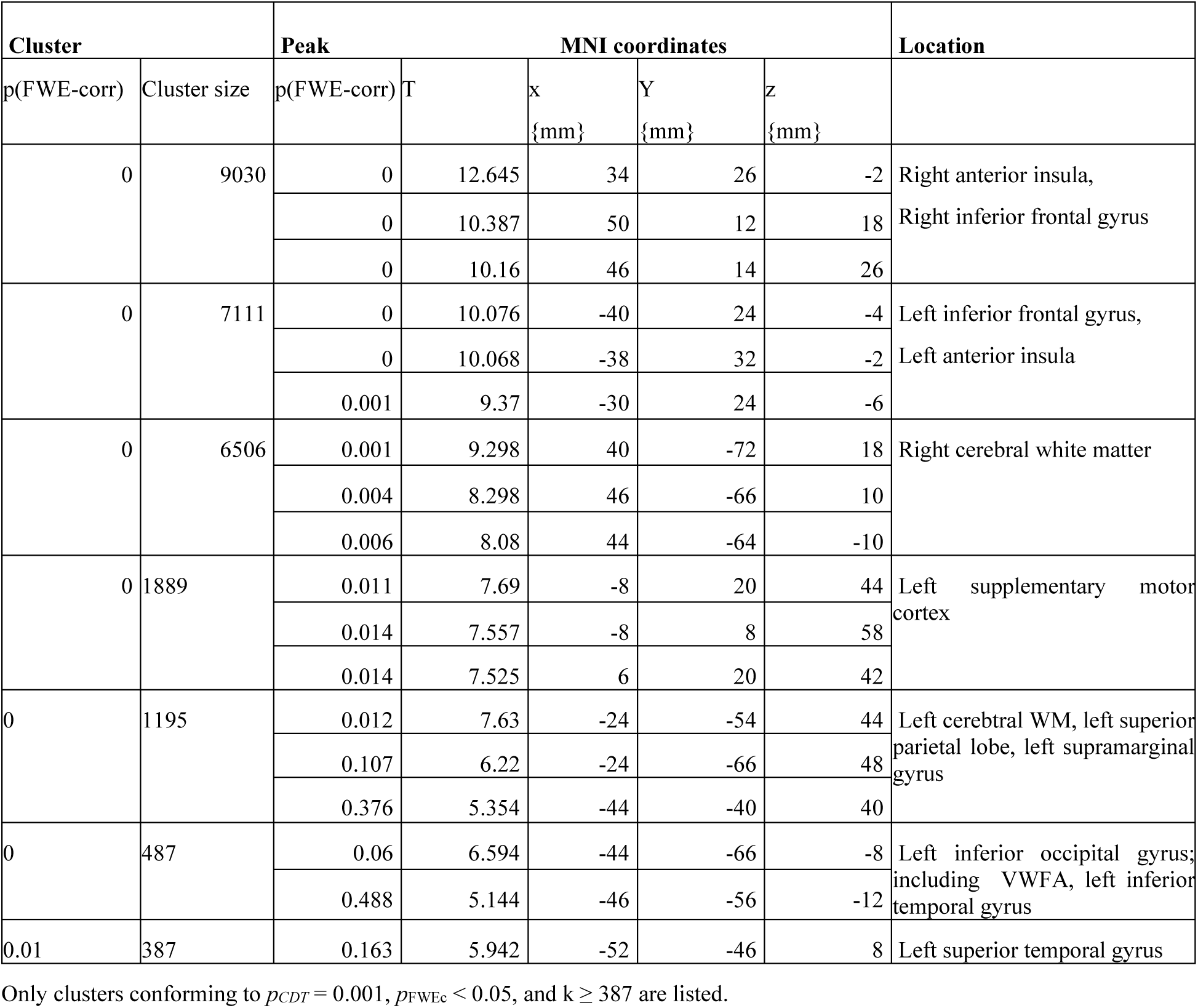
Significant activation clusters for the contrast regulation vs baseline averaged across six neurofeedback training runs for typical readers.

**Table 3.** Significant activation clusters for the contrast regulation vs baseline, averaged across six neurofeedback training runs for poor readers.

| Cluster |  | Peak |  |  |  |  | Location |
| --- | --- | --- | --- | --- | --- | --- | --- |
| p(FWE-corr) | Cluster size | p(FWE-corr) | T | x<br>{mm} | y<br>{mm} | z<br>{mm} |  |
| 0 | 23742 | 0 | 11.463 | 38 | 26 | -4 | Right inferior frontal gyrus |
|  |  | 0 | 10.925 | -40 | 24 | -4 |  |
|  |  | 0 | 10.62 | 50 | 18 | -8 |  |
| 0 | 2246 | 0.005 | 8.641 | 4 | 12 | 60 | Right supplementary motor cortex |
|  |  | 0.016 | 7.794 | 0 | 20 | 52 |  |
|  |  | 0.025 | 7.478 | 6 | 20 | 44 |  |
| 0 | 1139 | 0.106 | 6.471 | -30 | -64 | 54 | Left superior parietal lobe |
|  |  | 0.143 | 6.268 | -42 | -46 | 38 |  |
|  |  | 0.665 | 5.026 | -22 | -58 | 42 |  |
| 0.02 | 269 | 0.561 | 5.205 | 50 | -24 | -12 | Right middle temporal gyrus |
|  |  | 0.96 | 4.344 | 42 | -30 | -6 |  |
| 0.01 | 315 | 0.618 | 5.107 | -48 | -42 | 0 | Left middle temporal gyrus; including VWFA |
|  |  | 0.981 | 4.205 | -60 | -46 | 10 |  |
|  |  | 0.988 | 4.134 | -52 | -50 | 8 |  |

Only clusters, conforming to $p_{CDT} = 0.001$ , $p_{FWEc} < 0.05$ and $k \geq 0\ 315$ are listed.

### 3.4 Whole-brain Activation during No-feedback Runs Pre and Post Training

For no-feedback runs, we observed no significant differences between groups (typical and poor readers) and timepoints (pre and post neurofeedback training). For detailed information regarding the significant activation clusters identified for each group at different timepoints, please refer to Tables S6, S7, S8, and S9, as well as Figure S5 in the supplementary material. These supplementary materials provide a comprehensive overview of the activation patterns during the no-feedback runs observed in the study.

### 3.5 VWFA Activation and Time-course Analyses of Neuro- and No-feedback Runs

We used a mixed ANOVA to examine potential differences in the VWFA PSC across timepoints (before neurofeedback, during neurofeedback, after neurofeedback), groups (typical and poor readers), and upregulation phases during neurofeedback (first half of regulation block, second half of regulation block). This analysis revealed a main effect of timepoint (*F*(2, 72) = 21.5, p < 0.001), indicating a higher activation during the neurofeedback as compared to no-feedback runs. There was no main effect of phase (*F*(1, 36) = 2.17, p = 0.15) or group (*F*(1, 36) = 0.56, p = 0.5). In addition, we found a significant phase x timepoint interaction (*F*(2,72)= 8.91, *p* < 0.001) and a phase x group interaction on a trend level (*F(*1,36) = 3.81, p = 0.059). Post-hoc analyses showed a higher activation during neurofeedback compared to pre-training no-feedback runs in both early (*t*(379) = 8.51, p < 0.001) and late phases (*t*(379) = 14.7, *p* < 0.001). We also observed a higher activation during neurofeedback compared to post-training no-feedback runs in both phases, early (*t*(379) = 9.12, *p* < 0.001) and late (*t*(379) = 14.4, *p* < 0.001). Finally, we observed a higher activation during the post-training compared to pre-training no-feedback run (*t*(379) = 2.55, *p* = 0.01).

Parallel to MacInnes and colleagues (MacInnes et al. 2016), we performed one-sample t-tests against zero to investigate whether VWFA activation during different phases and time points significantly differed from baseline. Both groups showed significant positive activation in the early phase (TR: *t(*19) = 2.88, *p* = 0.009; PR: *t*(18)= 3.17, p = 0.005) as well as in the late phase (TR: *t*(19) = 3.39, p = 0.003; PR: *t*(18) = 3.86, *p* = 0.001) during neurofeedback training (Figure 7A). Individuals with typical reading skills also showed significant positive activation in the early phase of the post no-feedback run (*t*(18) = 2.94, *p* = 0.023), even though this result did not survive the correction for multiple comparisons. All other phases and timepoints did not significantly differ from the baseline. When performing group comparisons, we found no differences in VWFA activity between typical and poor readers for any phase or timepoint.

**Figure 7.**
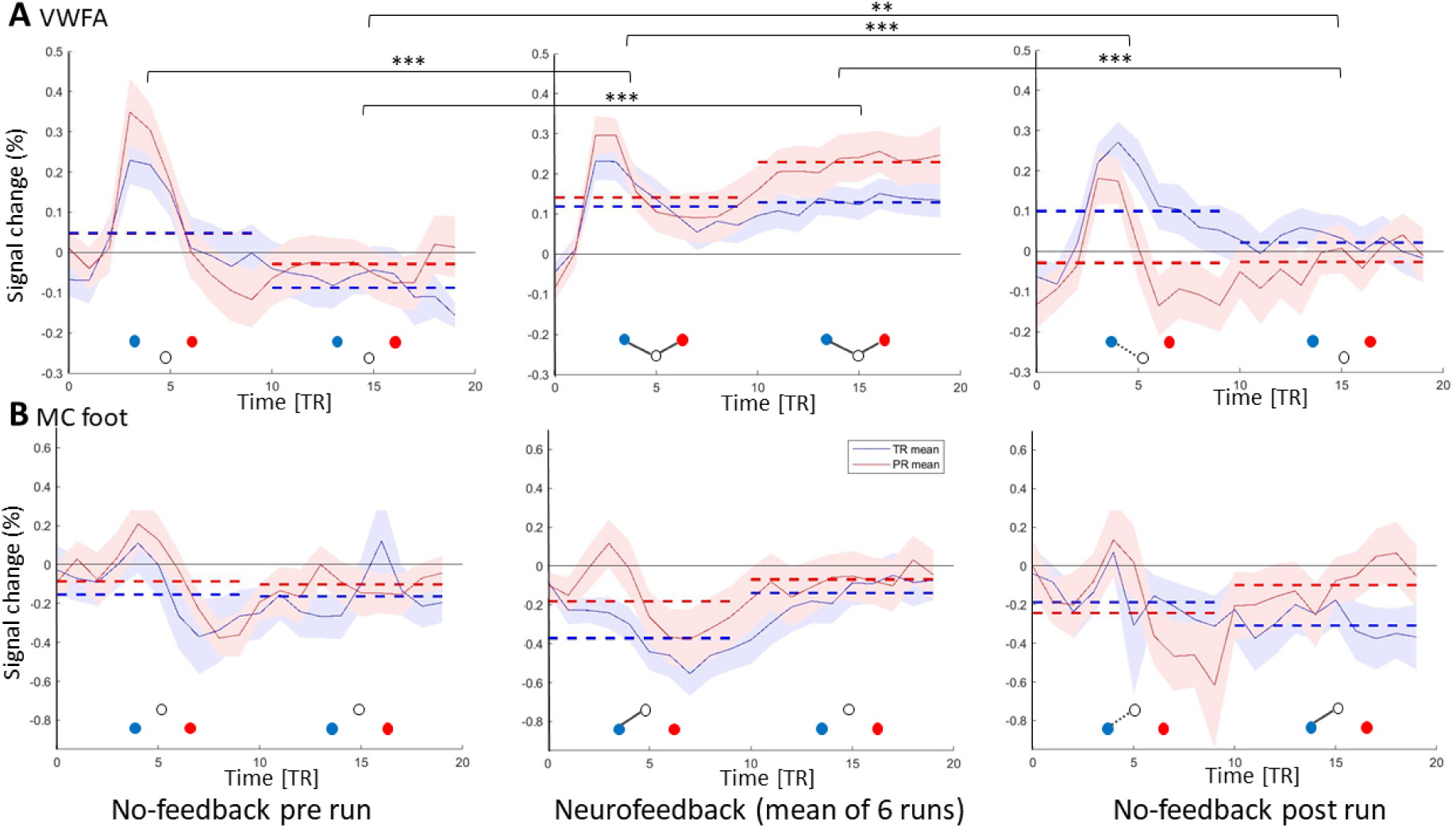
VWFA **(A)** or MC foot **(B)** activation time courses during pre-training no-feedback, neurofeedback (mean over six runs and averaged across the 4 blocks) and post-training no-feedback runs. The y-axis shows percent signal changes (regulation vs baseline) and the shaded areas indicate the corresponding standard deviation. Time courses were split into two halves of 20 seconds/10 TR’s (repetition time) each to examine differences in average PSC in the early and late phases of regulation. Dashed horizontal lines represent the means of each group in the two phases. The horizontal brackets indicate significant differences between the phases ** = p < 0.01, *** = p < 0.001 (FDR-corr. p < 0.05). As an example, the percent signal change during the late phase of no-feedback pre differs significantly from the signal change during the late phase of no-feedback post. Significant activation differences from baseline (FDR-corrected, p < 0.05) are indicated by black lines connecting the circles below the time courses. White circles represent the baseline level. When a white circle is positioned below the blue and red circles, it indicates values above zero (e.g. early and late phases of neurofeedback runs in VWFA, panel A) ; conversely, when the white circle is above the blue and red circles, it indicates values below zero (e.g. early phase of neurofeedback runs in MC foot, panel B). Examples for data interpretation: during neurofeedback training, PR (red) and TR (blue) showed significantly higher activation during regulation as compared with baseline in both phases, while this was not the case for the no-feedback runs. An example of a significant result not surviving multiple comparisons is the VWFA post no-feedback run in readers with typical reading skills. Blue circle = typical readers, red circle = poor readers, black lines = significant results FDR corrected for multiple comparisons (FDR-corr. p < 0.05), dotted black line = p<0.05, uncorrected.

Finally, we repeated our analyses for other regions of the reading network as well as a control region unrelated to reading (MC foot). For the MC foot ROI, we observed a significant deactivation during the early phase of neurofeedback runs for individuals with typical reading skills (TR: *t*(19) = −4.40, *p* = 0.0003). A visual presentation of the time courses for the additional ROIs of the reading network (including the IFG, IPL, PrecG, and STG) is provided in supplementary material Figure S6.

### 3.6 Mental Strategies during the Neuro- and No-feedback Training

Participants with typical reading skills used the *“imagine reading”* strategy significantly more often than those with poor reading skills during neurofeedback runs (*χ2*(1) = 10.36, *p* = 0.005) (Figure 8A, Table 1). During the post-training no-feedback run, participants with poor reading skills did not show a significant difference in the use of “*imagine reading”* strategies compared to those with typical reading skills (*χ2*(1) = 1.51, *p* = 0.273) (Figure 8B, Table 1). The visualization of the use of the strategies (based on the six categories) is presented in supplementary material Figure S7.

**Figure 8.**
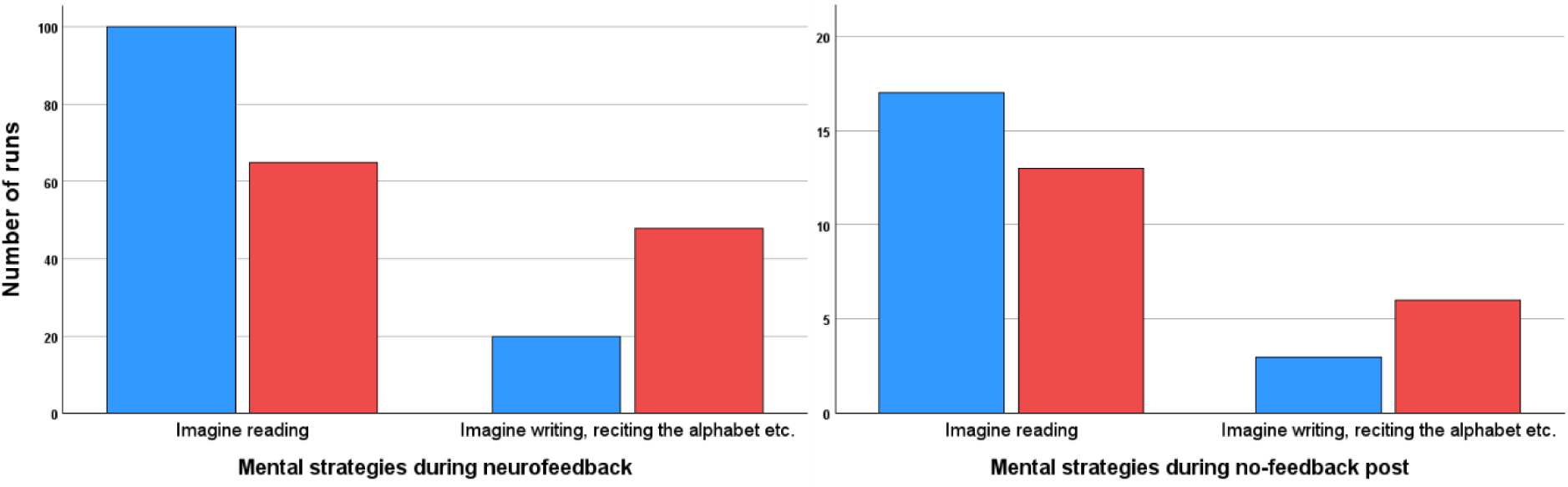
A. Strategies chosen over the six neurofeedback runs. Participants with typical reading skills demonstrated a significantly higher preference for imagined reading strategies compared to those with poor reading skills (χ2((1) = 10.365, p < 0.005) throughout the neurofeedback sessions. **B. Strategies chosen for the post no-feedback run.** There was no statistically significant difference between the two groups in the post no-feedback run (χ2((1) = 1.509, p < 0.273) concerning employing the imagine reading strategy. Typical readers are represented in blue,while poor readers are represented in red.

Moreover, there was no significant difference in the number of strategies employed during the neurofeedback sessions between the two groups (*t*(37) = −0.009, *p* = 0.874) (number of strategies mean = 2.13, range [1-4]. See Table 1).

## 4. Discussion

This study aimed to investigate the feasibility of using real-time fMRI neurofeedback to modulate neural activity in the VWFA across the full spectrum of reading competence, with a particular emphasis on individuals at the lower end of the spectrum. Building on previous research demonstrating reduced VWFA activation in individuals with poor reading skills (e.g. (Dehaene and Cohen 2011; Brem et al. 2010; Richlan et al. 2009)) our study examined whether VWFA activity could be modulated using rt-fMRI neurofeedback. Our main findings, detailed in the following chapters, are as follows: First, participants with both typical and poor reading skills initially struggled to activate the VWFA using reading-related mental strategies during the pre-training no-feedback run. However, with rt-fMRI neurofeedback, both groups were able to achieve voluntary VWFA activation. Second, post-training, sustained VWFA activity during the no-feedback run was significantly elevated compared to pre-training levels across groups, regardless of reading proficiency. Notably, VWFA activation during neurofeedback sessions was substantially higher than during no-feedback runs. Finally, individuals with poor reading skills reported using imagined reading strategies less frequently during neurofeedback training than those with typical reading skills. These results demonstrate the potential of rt-fMRI neurofeedback to facilitate VWFA self-regulation across a wide range of reading abilities.

### 4.1 Reduced Activity in the VWFA with Poor Reading Fluency

It is well-established, as supported by meta-analyses (Richlan et al. 2011, 2009; Martin et al. 2016), that aberrant function in the VWFA is associated with impaired reading skills and limited reading experience. Dehaene et al. (Dehaene et al. 2010), for example, observed a correlation between the strength of VWFA responses during reading tasks and the level of reading expertise in illiterate, ex-illiterate, and literate adults. Similarly, a multi-centre neuroimaging study (Brem et al. 2020) found that word reading fluency strongly influenced VWFA activity in children, with the association being particularly pronounced in those with lower reading abilities. Our findings from the functional VWFA localizer align with these studies, demonstrating a significant positive correlation between pseudoword reading fluency and VWFA activity. In our adult group, reduced word-sensitive activation in the VWFA during word processing was associated with poor reading skills. This aberrant functional activity underscores the VWFA’s potential as a target for rt-fMRI-based interventions aimed at improving reading skills and serves as the foundation for our neurofeedback trial. Besides replicating the link between reading competence and VWFA activation, we also used our functional VWFA localizer task to determine each individual’s VWFA for the neurofeedback approach. This is crucial given the variability in VWFA location across individuals (Glezer et al. 2009; Pleisch et al. 2019; Dehaene-Lambertz et al. 2018). With our localizer task, we successfully detected VWFA activity at the single-subject level in both groups: adults with typical and poor reading skills. A reliable localizer task that can detect VWFA activity regardless of reading skills is essential for neurofeedback training to target normalisation of VWFA in individuals with poor reading skills.

### 4.2 Upregulation of the VWFA with Feedback in Real-time

Our results demonstrate that adult participants, regardless of their reading skills, successfully learned to upregulate VWFA activity during neurofeedback training. FMRI analyses revealed significant activation in the VWFA during neurofeedback for both groups. ROI analyses of the VWFA furthermore showed significant activation levels above zero during both the early and late phases of neurofeedback training blocks in both groups.

Our findings build upon and extend our previous work (Haugg et al. 2023), using the same group of participants with typical reading skills to another cohort of typical readers instructed to downregulate VWFA activity using the identical experimental design. In this previous study, we observed a significant difference in VWFA activation during neurofeedback runs between the downregulation and upregulation groups, both of which received real-time feedback on VWFA activity. Expanding upon these findings, we demonstrate that VWFA activity can be modulated regardless of reading proficiency, as even adults with poor reading skills can learn to upregulate this region. This aligns with numerous studies reporting successful regulation of targeted brain areas across a range of neurological (e.g (Emmert et al. 2017; Papoutsi et al. 2020; Robineau et al. 2019; Buyukturkoglu et al. 2013)) and psychiatric disorders (Stoeckel et al. 2014; Tursic et al. 2020). These findings span various subcortical and cortical brain regions, including the temporal cortex (Pereira et al. 2019; Habes et al. 2016). Also, individuals with Autism Spectrum Disorder (ASD) have successfully learned to self-regulate activity in key brain areas, including the insula, fusiform face area, and posterior superior temporal sulcus (Caria et al. 2012; Pereira et al. 2019; Caria and Falco 2015; Direito et al. 2021). Similarly, in individuals with Attention-Deficit/Hyperactivity Disorder (ADHD), rt-fMRI neurofeedback has been successfully used to modulate activity in the right inferior prefrontal cortex, a region critical for attention and executive function (Alegria et al. 2017; Rubia et al. 2019).

Our findings also show that during neurofeedback training, the measured activation was significantly higher than in the pre- and post-training no-feedback runs, both in the early and late phases of regulation blocks, across both groups. This highlights the importance of feedback for learning to upregulate the VWFA. While reading is a complex cognitive process, and the VWFA operates as part of an intricate network, making it challenging to align feedback with the precise neural activity (compare (Lubianiker et al. 2022)), our participants were nonetheless able to effectively regulate VWFA activity. This finding indicates that real-time feedback can be an effective tool for enabling individuals to voluntarily modulate activity in specific brain regions, even during complex cognitive tasks such as mentally imagining reading.

The result that participants with and without poor reading skills were capable of upregulating VWFA activity is especially noteworthy, given that individuals with poor reading skills typically exhibit reduced VWFA responses to written stimuli (Dehaene and Cohen 2011; Dehaene et al. 2010; Brem et al. 2010), a pattern that was also observed with our VWFA localizer task. These results imply that, although reading tasks or viewing written stimuli often fail to elicit comparable activation, mental imagery of reading, when reinforced by feedback, can evoke similar levels of VWFA activity between participants with typical and poor reading skills. Thus, rt-fMRI feedback likely plays a crucial role in guiding participants to discover suitable mental and neural strategies that enhance VWFA activation. This finding is noteworthy because it suggests that mental imagery of reading, when combined with rt-fMRI neurofeedback, can effectively engage brain areas involved in reading. This approach could offer a novel strategy for further exploration in supporting interventions for individuals with reading impairments.

#### Comprehensive Findings on Reading Network Activation

Furthermore, both groups, regardless of their reading skills, showed activation in additional core regions of the reading network during neurofeedback training. This suggests that the reading-related imagery task, supported by real-time feedback of brain activation, engages a broader neural reading-related network. Specifically, the activated regions included the left IFG, precentral gyrus (PreG), left angular gyrus, supramarginal gyrus, and STG. This network, known as the reading network (Turker et al. 2025), is involved in various functions to process written information, including phonological processing (Costafreda et al. 2006), semantic integration (Horwitz et al. 1998), LSS integration (van Atteveldt et al. 2004), motor articulation (Baldo et al. 2011), and auditory comprehension (Buchsbaum et al. 2001; Hickok et al. 2000; Yi et al. 2019; van Atteveldt et al. 2004). The left IFG is involved in semantic processing (Carota et al., 2017) and LSS mapping (Sandak et al., 2004), and it has strong reciprocal connections to the left occipitotemporal cortex (Costafreda et al. 2006). The Precentral Gyrus (PreG) plays a critical role in coordinating the movements necessary for speech production (Baldo et al. 2011). Activation of these frontal regions during neurofeedback training thus suggests that participants may use motor imagery and subvocal articulatory rehearsal strategies to facilitate the regulation of VWFA. The STG is involved in auditory processing and language comprehension (Buchsbaum et al. 2001; Yi et al. 2019) and thus may indicate that participants engage in higher-level auditory processing and language comprehension evoked by their mental imagery. Finally, the left angular and supramarginal gyri play a key role in various complex language functions, including multimodal integration, attention, reading, writing, and comprehension (Horwitz et al. 1998; Kearns et al. 2019). The activation pattern thus suggests that participants are engaging in lexico-semantic processing and integrating multimodal information to improve their reading-related mental strategies.

On the one hand, the co-activation of those regions demonstrates the effectiveness of the training, as it enabled the activation of the entire network through imagination aided by feedback, a phenomenon not observed before the training. On the other hand, it highlights the complexity of VWFA activation as the region is part of an intricate network involved in reading processing. Moreover, this broader activation supports the idea that neurofeedback not only targets specific brain regions but also facilitates the integration and optimisation of related neural circuits, thereby enhancing overall cognitive function related to the task at hand. This finding is consistent with other neurofeedback studies, which have demonstrated that the impact of neurofeedback can extend beyond the targeted brain region. For instance, some researchers (Rubia et al. 2019; MacInnes et al. 2016) found that neurofeedback modulated connectivity throughout the entire mesolimbic network. Furthermore, Zweerings and colleagues (Zweerings et al. 2019) observed changes in resting state connectivity networks after the successful training of core language network nodes using neurofeedback.

Moreover, many of these regions would be valuable targets for inclusion in neurofeedback training, as they have been reported to be hypoactive during reading tasks in individuals with reading disorders (Martin et al. 2016; Yan et al. 2021; Richlan et al. 2009). Next, to train these regions separately, future studies should consider employing connectivity-based neurofeedback approaches, which enable the modulation of functional connectivity between targeted brain areas (e.g., (Koush et al. 2013; Morgenroth et al. 2020; Liew et al. 2016).

Since whole-brain activation during neurofeedback showed no group differences between individuals with typical reading skills and poor reading skills, we can infer that participants with poor reading skills utilize a similar level and pattern of reading network activity during upregulation of the VWFA.

### 4.2 Upregulation without Feedback: No-feedback runs pre and post

To investigate the regulation performance without providing feedback, we performed two no-feedback runs, before and after neurofeedback training. Our results show no VWFA activation in the no-feedback run before the training. Mental imagery of reading alone appears insufficient to reliably elicit a VWFA response, regardless of reading skill level. This result is consistent with the findings of MacInnes et al (MacInnes et al. 2016), demonstrating that participants were initially unable to increase activation in targeted brain areas (VTA, NAcc) using mental strategies alone before the neurofeedback training. Regulation was only successful when they received neurofeedback from the target regions. Even though there is evidence that other brain regions, such as the FFA, PPA, and the LOC (Reddy et al. 2010), can be activated by imagining objects similar to visual presentation, this seems not to be the case in the VWFA and with reading. Reading is a complex process, and developing effective strategies to regulate VWFA activity is particularly challenging, especially since brain activity is not directly accessible to conscious awareness. Nevertheless, becoming aware of neural activity changes during regulation attempts through feedback appears to facilitate the learning process (Muñoz-Moldes and Cleeremans 2020; Lacroix and Gowen 1981). Furthermore, the VWFA is part of the broader and intricate reading network. Feedback about the neuronal activation level of the VWFA appears to be crucial for participants to explore and develop effective strategies to upregulate VWFA activity.

In the post no-feedback run, whole brain analyses showed no group difference between participants with poor reading skills and those with typical reading skills.

Time-course analyses of percentage signal change (PSC) revealed that the increase in VWFA activation observed across groups during neurofeedback training was due to a significant difference in average activity during neurofeedback runs compared to no-feedback runs, both before and after training, and in both early and late phases. The higher activation observed in the post-training run compared to the pre-training run suggests a reactivation of neural states learned during neurofeedback. This indicates that, following neurofeedback training, participants are better able to regulate VWFA activation even without feedback, regardless of their reading skills, highlighting the potential of neurofeedback to facilitate self-regulation of brain activity.

Many fMRI neurofeedback studies have reported lasting effects of neurofeedback on behaviour or activity (Young et al. 2017; MacInnes et al. 2016). However, our effects appear to be rather modest. As previously mentioned, while we observed overall higher PSC in the post no-feedback runs compared to pre no-feedback runs, this elevated PSC level was not sustained (not significantly different from baseline activity in the late phase). This contrasts with the findings of MacInnes and colleagues (MacInnes et al. 2016), who reported that healthy participants receiving feedback from the targeted brain regions showed significantly higher activation in those regions during both neurofeedback and subsequent no-feedback runs, compared to participants who received feedback from the alternate regions. Furthermore, their study observed sustained activation in the late phase of the post no-feedback run in the feedback group, a pattern not evident in the two control groups (false feedback and feedback from alternate regions). This raises the question of why it is particularly challenging to maintain VWFA activity without ongoing feedback. Our relatively lengthy training protocol (six runs) and the scheduling of the post-run within the same session may have contributed to enhanced neural adaptation effects (e.g. (Perrachione et al. 2016)). However, this design could also have resulted in increased fatigue and reduced vividness of mental imagery. Additionally, because the VWFA operates within a complex neural network and its activation is not consciously accessible, sustaining its activity without feedback may remain a significant challenge.

### 4.3 Mental Strategies Applied During Upregulation Training

Our results indicate that adults with poor reading skills were less likely to choose reading-related mental strategies during regulation. However, they did not change strategies more frequently, suggesting they may not have been fully aware that their chosen approaches were less related to reading. The choice of mental strategies in neurofeedback training requires careful consideration. While instructing participants to use a specific strategy, such as imagining letter strings, could help standardize methods across individuals, it may also have drawbacks. Overly prescriptive instructions might limit participants’ ability to explore and identify more effective, personalized strategies (Hardman et al. 1997). This consideration is particularly crucial for participants with learning disorders. For these individuals, the use of compensatory strategies presents a complex challenge: while such strategies can help leverage existing strengths, they may also reinforce less optimal neural pathways. As a result, decisions about encouraging or discouraging compensatory strategies during neurofeedback training should be made thoughtfully, taking into account each individual’s needs and training objectives. Ultimately, the most effective approach may strike a balance between standardized methods and the flexibility to allow for personalized exploration.

Despite the variety of strategies employed, the impact of the different strategies on participants’ ability to upregulate VWFA activity was not statistically significant. This lack of clear superiority among different mental approaches aligns with observations from EEG-based neurofeedback studies, highlighting a common challenge in neurofeedback research. Autenrieth et al. (Autenrieth et al. 2020) and Hardman et al. (Hardman et al. 1997) reported no evidence that specific mental strategies are more effective than others, particularly in single-session neurofeedback studies, where direct comparisons are inherently challenging. In our study, participants completed six neurofeedback runs, offering substantial opportunity to refine and evaluate their mental strategies across sessions. Despite this extended exposure, we found no significant correlation between the type of strategy chosen and participants’ success in upregulating VWFA activity, regardless of whether they had typical or poor reading skills. This suggests that simply having more time to experiment with or optimize strategies does not necessarily translate into better neurofeedback outcomes, a finding consistent with broader neurofeedback literature, where the efficacy of specific mental strategies remains inconclusive even after multiple sessions (Kober et al. 2013; Haugg et al. 2021; Thibault et al. 2015). Nevertheless, providing careful instruction and guidance in strategy selection remains essential. Such support can help participants avoid counterproductive approaches, enhance cognitive engagement, and facilitate more effective regulation of the target brain region during neurofeedback training (Emmert et al. 2017; Lubianiker et al. 2019).

Our study is consistent with previous research utilizing explicit feedback processing, where participants receive direct instructions to employ specific mental strategies for regulating targeted brain activity, and are subsequently provided with visual feedback reflecting their success (Auer et al. 2015). In contrast, other studies adopt an implicit feedback processing approach, withholding explicit instructions and allowing participants to learn how to modulate brain activity without conscious awareness (e.g. (Amano et al. 2016; Taschereau-Dumouchel et al. 2021)). The relative effectiveness of explicit versus implicit feedback processing can vary, and the optimal choice may depend on the objectives and context of the neurofeedback intervention, as well as the characteristics of the participant population (Lubianiker et al. 2019; Watanabe et al. 2017; Sitaram et al. 2024).

Researchers continue to debate whether neurofeedback training primarily relies on subconscious brain regulation or is driven by deliberate, intentional strategies. Some evidence suggests that neurofeedback is most effective when participants engage in volitional self-regulation using consciously selected mental strategies, supporting a top-down, cognitively controlled process (e.g., (Sitaram et al. 2017; Sitaram et al. 2024; Bagdasaryan and van Quyen 2013). Conversely, other studies indicate that successful regulation can occur even without conscious effort, pointing to bottom-up, automatic mechanisms that may operate outside of conscious awareness (e.g., (Sitaram et al. 2024; Watanabe et al. 2017; Shibata et al. 2019; Oblak et al. 2019)). Despite these differing viewpoints, few empirical studies (e.g., (Sepulveda et al. 2016)) have explicitly examined these two hypotheses, leaving the debate largely unresolved.

Future research should investigate how best to balance structured strategy instruction with opportunities for participants to independently discover effective approaches, as both methods may offer unique benefits depending on individual learning styles and neurofeedback goals. Additionally, examining the long-term outcomes of neurofeedback training, particularly its durability and transferability, as well as assessing its effectiveness in clinical populations, such as individuals with learning disorders or reading difficulties, remains a critical direction for advancing the field.

### 4.5 Limitations and Outlook

This study demonstrates that rt-fMRI neurofeedback can feasibly be used to upregulate VWFA activity in adults with a range of reading abilities. However, we did not assess whether this neural modulation translates into measurable improvements in reading skills. The short training duration and the focus on adults, whose reading networks are already well-established, limit the ability to evaluate the direct impact of neurofeedback on reading performance. As a result, while our findings are promising from a neurobiological perspective, they do not provide evidence for enhanced reading ability.

Future research should consider targeting younger learners, who may benefit more from VWFA upregulation due to greater neural plasticity during the critical period of reading acquisition. Extending the number of neurofeedback sessions, increasing the overall training duration, and incorporating follow-up assessments would help clarify the long-term effects of VWFA-targeted neurofeedback on reading skills. Additionally, our relatively small sample size, while comparable to prior neurofeedback studies that detected significant effects, may have limited our ability to identify more subtle improvements. Larger-scale studies are warranted to more robustly evaluate the efficacy of this approach for enhancing reading outcomes (e.g. (MacInnes et al. 2016; Pereira et al. 2019; Pamplona et al. 2020; Watve et al. 2024; Haugg et al. 2023; Haugg et al. 2021)).

### 4.6 Conclusion

This neurofeedback study offers two key insights: First, it demonstrates that adults with poor reading skills are capable of upregulating VWFA activity using rt-fMRI neurofeedback. Second, the intervention effectively increased VWFA activation even during a subsequent transfer run without real-time feedback, indicating that the effects of neurofeedback training can persist beyond the immediate feedback context. This enhancement was observed in both typical and poor readers, suggesting the potential for durable changes in brain function following VWFA-targeted neurofeedback. Notably, individuals with poor reading skills, despite initially showing lower VWFA activity during the localizer task and a tendency to use less “imagine reading” as a mental strategy, were still able to learn to regulate their VWFA activity through neurofeedback. These findings establish a promising basis for future interventions designed to influence reading network activity in individuals with reading difficulties, with the ultimate goal of enhancing reading skills.

## Data availability

All code used for MRI data analyses can be found on our Open Science Framework repository: https://osf.io/w3xzv/. The repository also includes exam cards with detailed information on MR parameters and the literature-based mask of the VWFA. Data will be made available upon written request to the corresponding author. A formal data sharing agreement will be required.

## Acknowledgment

We are grateful for the support of M. Röthlisberger in creating the localizer task stimulus, and to Milena Menghini, Felizia Stutz, and Sara Steinegger for their contribution to behavioural assessments and MRI recordings. Finally, we thank all participants for participating in this study.

## Declaration of generative AI in scientific writing

The authors declare the use of AI (Grammarly, Chat-GPT-4, and Perplexity) in the writing process to enhance grammar, sentence structure, wording, and language of the manuscript.

## Ethics Approval and Clinical Trial Registration

The trial was registered prospectively at the Swiss National Clinical Trial Portal (kofam.ch) with the identifier SNCTP000004276. The project received approval from the local ethics committee of the Canton of Zurich and adhered to the guidelines outlined in the Declaration of Helsinki (BASEC number 2021-00071).

## CRediT authorship contribution statement

Nada Frei: Conceptualization, Methodology, Formal analysis, Investigation, Writing-Original Draft, Writing-Review & Editing, Visualisation, Supervision, Project administration

Amelie Haugg: Conceptualization, Methodology, Investigation, Writing-Original Draft, Writing-Review & Editing, Supervision, Project administration

Silvia Brem: Conceptualization, Resources, Writing-Original Draft, Writing-Review & Editing, Funding acquisition

Gustavo Pamplona: Formal Analysis, Writing-Review & Editing, Visualisation

