## Supplementary Material for "Regulating brain activity in the Visual Word Form Area with real-time fMRI neurofeedback in adults with typical and poor reading skills"

#### 1. Methods

##### 1.1 Literature-based Visual Word Form Area Mask

The same VWFA mask as in our previous study, with some of the same participants (Haugg et al. 2023) was used. This VWFA is based on literature findings and was generated by delineating spheres with varying radii around the reported activation peaks from the listed articles on VWFA. Subsequently, these spherical Regions of Interest (ROIs) were combined to create a unified VWFA mask using MarsBaR. This unified mask was then used to define individual ROIs for neurofeedback training by selecting the 50% most active voxels within the mask, which served as the basis for providing feedback on the BOLD signal.

**Table S1. Literature of the visual word form area (VWFA) coordinates**

| **Literature** | **Coordinates (x,y,z in MNI space)** | **Radius** |
|---|---|---|
| *Impact of literacy on the functional connectivity of vision and language-related networks (López-Barroso et al. 2020)* | -44, -50, -14 | 8 mm |
| *Brain sensitivity to print emerges when children learn letter–speech sound correspondences (Brem et al. 2010)* | -48, −66, −14; | 6 mm |
| *Variability in Location Impacts Orthographic Selectivity in the "Visual Word Form Area" (Glezer and Riesenhuber 2013)* | -45, -56, -16 | 4 mm |
| *Lateralized task shift effects in Broca's and Wernicke's regions and the visual word form area are selective for conceptual content and reflect trial history (Wallentin et al. 2014)* | -43, -54, -12 | 10 mm |
| *The visual word form area and the frequency with which words are encountered: evidence from a parametric fMRI study (Kronbichler et al. 2004)* | -43, -54, -12 | 3 mm |
| *The Putative Visual Word Form Area Is Functionally Connected to the Dorsal Attention Network (Vogel et al. 2012)* | -45, -57, -12 | 4 mm |
| *The VWFA Is the Home of Orthographic Learning When Houses Are Used as Letters (Martin et al. 2019)* | -34, -55, -13 | 6 mm |
| *Converging evidence for functional and structural segregation within the left ventral occipitotemporal cortex in reading (Lerma-Usabiaga et al. 2018)* | -42, -58, -10 | 6 mm |

Coordinates and radii of the spheres were chosen based on the literature. A general visual word form area mask was created based on previous literature.

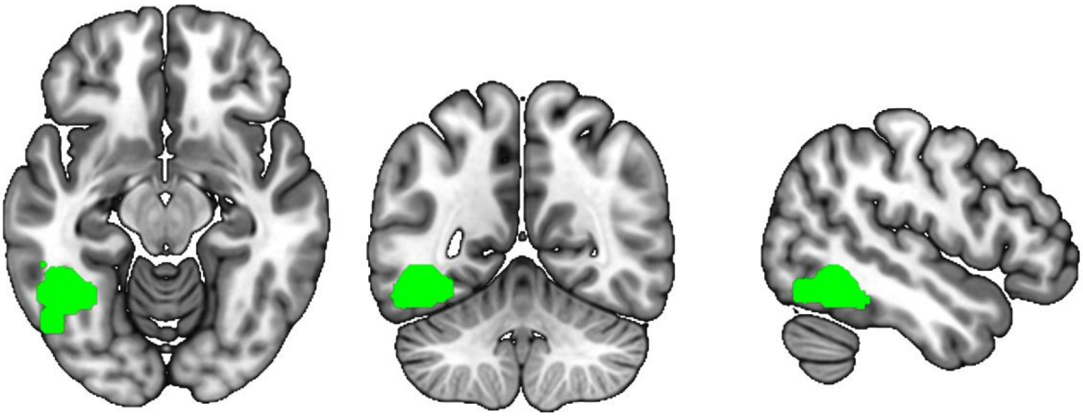

**Figure S1. Literature-based mask of the Visual Word Form Area (VWFA).** The literature-based VWFA mask was constructed by combining spheres of varying radii, with coordinates and radii derived from published studies (see Table S1).

#### 1.2 Task Design No-feedback

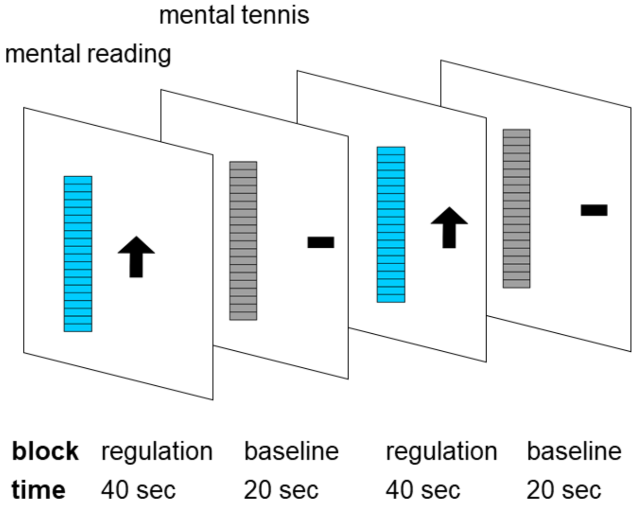


**Figure S2. No-feedback paradigm.** A blue static thermometer is presented during the regulation blocks and a grey static thermometer during rest blocks. Duration and instructions were the same as for the neurofeedback runs.

#### 1.3 Rating of used Mental Strategies during Regulation Training

**Table S2. Mental strategies during no- and neurofeedback regulation**

| Mental imagery of: | Imagine reading |
|---|---|
| Reading text | 1 |
| Reading words | 1 |
| Reading letters | 1 |
| Writing | 0 |
| Spoken language | 0 |
| Others | 0 |

Eleven independent team members evaluated the strategies used during the regulation blocks of both the no-feedback and neurofeedback runs, categorizing them into six distinct subcategories. For analysis, these categories were further consolidated into two main categories: 'imagine reading' (1) and 'not imagine reading.' (0).

### 2. Results

#### 2.1 Localizer Run

##### 2.1.1 Single Subject Level

All participants with typical reading skills exhibited significant VWFA activation on whole-brain analysis during the pre-training localizer run (initial threshold $p_{CDT} = 0.001$, $p_{\text{FWEc}} < 0.05$). Only one participant with poor reading skills showed significant VWFA activation during the pre-training localizer run (initial threshold $p_{CDT} = 0.001$, $p_{\text{FWEc}} < 0.05$). However, among participants with poor reading skills, all but one showed significant VWFA activation using small volume correction.

**Table S3. Single-subject level VWFA activation (beta values) of typical and poor readers**

| Typical readers | | | | Poor readers | | | | |
|---|---|---|---|---|---|---|---|---|
| | **FWE corrected cluster** $p_{\text{FWEc}}$ | **Initial threshold** $p_{CDT}$ | **VWFA beta values** | | **FWE corrected cluster** $p_{\text{FWEc}}$ | **Initial T Threshold** $p_{CDT}$ | **VWFA beta values** | **Significant cluster in VWFA (Small Volume )** |
| TR_01 | p < 0.001 | 0.001 | 0.5597 | PR_01 | - | 0.001 | 0.5221 | 0.044 |
| TR_02 | p < 0.001 | 0.001 | 0.6589 | PR_02 | - | 0.001 | 0.735 | 0.044 |
| TR_03 | p < 0.001 | 0.001 | 0.797 | PR_03 | - | 0.001 | 0.7529 | < 0.001 |
| TR_04 | p < 0.001 | 0.001 | 0.4267 | PR_04 | 0.534 | 0.001 | 0.3385 | 0.007 |
| TR_05 | p < 0.001 | 0.001 | 0.5946 | PR_05 | | 0.001 | 0.4234 | < 0.000 |
| TR_06 | p < 0.001 | 0.001 | 0.1386 | PR_06 | | 0.001 | 0.2145 | 0.019 |
| TR_07 | p < 0.001 | 0.001 | 0.1373 | PR_07 | - | 0.001 | 0.2489 | 0.001 |
| TR_08 | p < 0.001 | 0.001 | 0.417 | PR 08 | - | 0.001 | 0.6437 | <0.001 |
| TR_09 | p < 0.001 | 0.001 | 0.4393 | PR_09 | - | 0.001 | 0.3686 | <0.001 |
| TR_10 | p < 0.001 | 0.001 | 0.3749 | PR_10 | - | 0.001 | 0.8501 | <0.001 |
| TR_11 | p < 0.001 | 0.001 | 0.5523 | PR_11 | - | 0.001 | 0.4252 | 0.008 |
| TR_12 | p < 0.001 | 0.001 | 0.5799 | PR_12 | < 0.001 | 0.001 | 0.3903 | <0.001 |
| TR_13 | p < 0.001 | 0.001 | 0.1715 | PR_13 | - | 0.001 | 0.3011 | - |
| TR_14 | p < 0.001 | 0.001 | 0.4536 | PR_14 | - | 0.001 | 0.7482 | <0.001 |
| TR_15 | p < 0.001 | 0.001 | 0.5536 | PR_15 | - | 0.001 | 0.5312 | <0.001 |
| TR_16 | p < 0.001 | 0.001 | 0.3087 | PR_16 | - | 0.001 | 0.3167 | 0.001 |
| TR_17 | p < 0.001 | 0.001 | 0.675 | PR_17 | - | 0.001 | 0.0056 | <0.001 |
| TR_18 | p < 0.001 | 0.001 | 0.3006 | PR_18 | | 0.001 | 0.578 | <0.001 |
| TR_19 | p < 0.001 | 0.001 | 1.0926 | PR_19 | - | 0.001 | 0.24282 | <0.001 |
| TR_20 | p < 0.001 | 0.001 | 0.4614 | | | | | |

#### 2.1.2 Correlation Pseudoword Reading Fluency and VWFA Activity

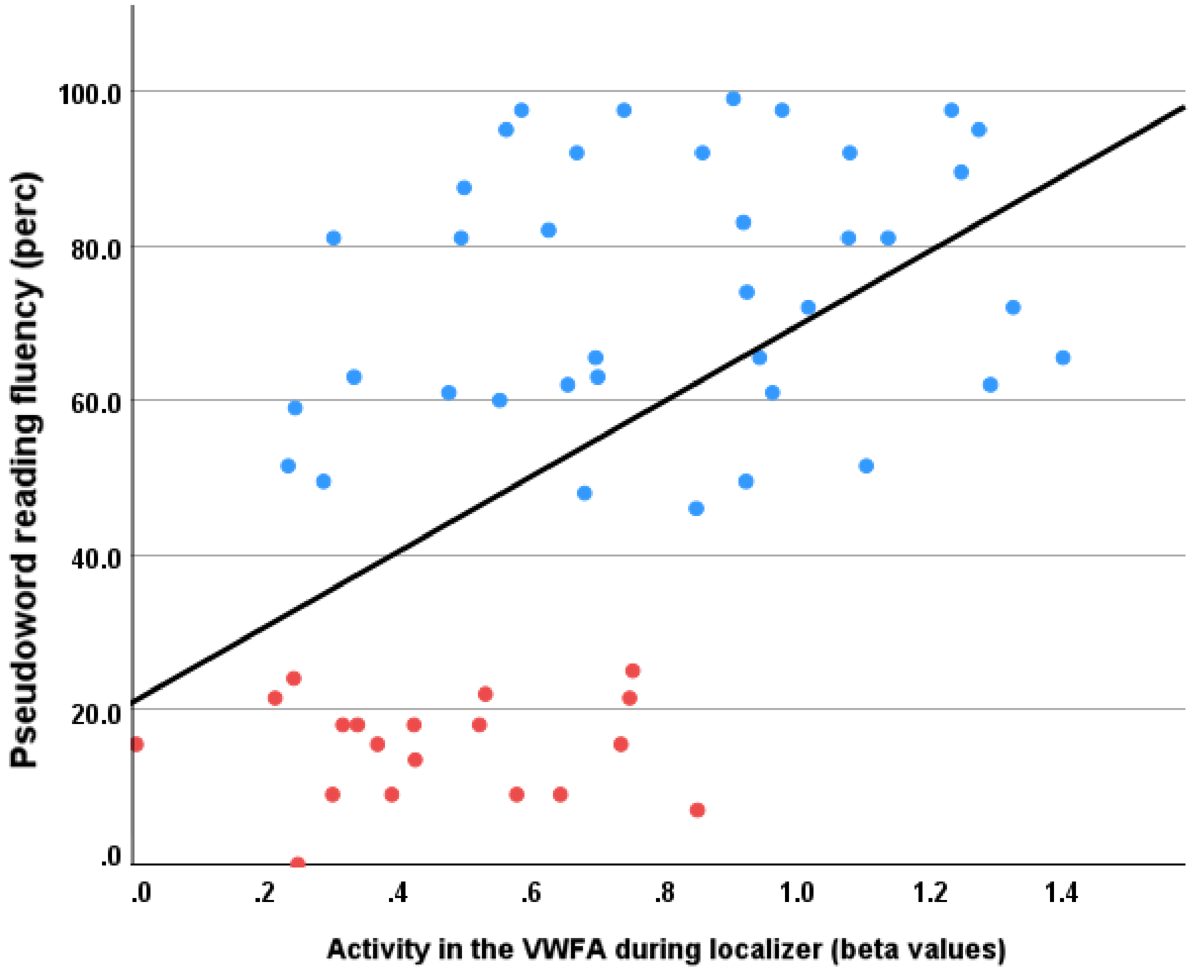


**Figure S3.** A significant positive correlation was observed between pseudoword reading fluency percent scores (SLRT-II) and VWFA beta values for the words vs. checkerboards contrast in an enlarged sample ($r = 0.523$, $p < 0.001$, $n = 57$), after adding 18 typical readers from our previous study (Haugg et al. 2023) to the original sample. Participants with typical reading skills are shown in blue, while those with poor reading skills are shown in red.

#### 2.1.3 Localizer Activity in the Groups of Typical and Poor Readers

The group of participants with typical reading skills showed significant activation in bilateral inferior occipital gyri, left inferior frontal gyrus, left superior parietal lobe, right pre- and post-central gyri, supplementary motor cortex, and left middle temporal gyrus.

The group of participants with poor reading skills exhibited significant activity in the bilateral inferior occipital gyri, the left precentral gyrus, left angular gyrus, right precentral gyrus, right middle frontal gyrus (MFG), and right inferior frontal gyrus.

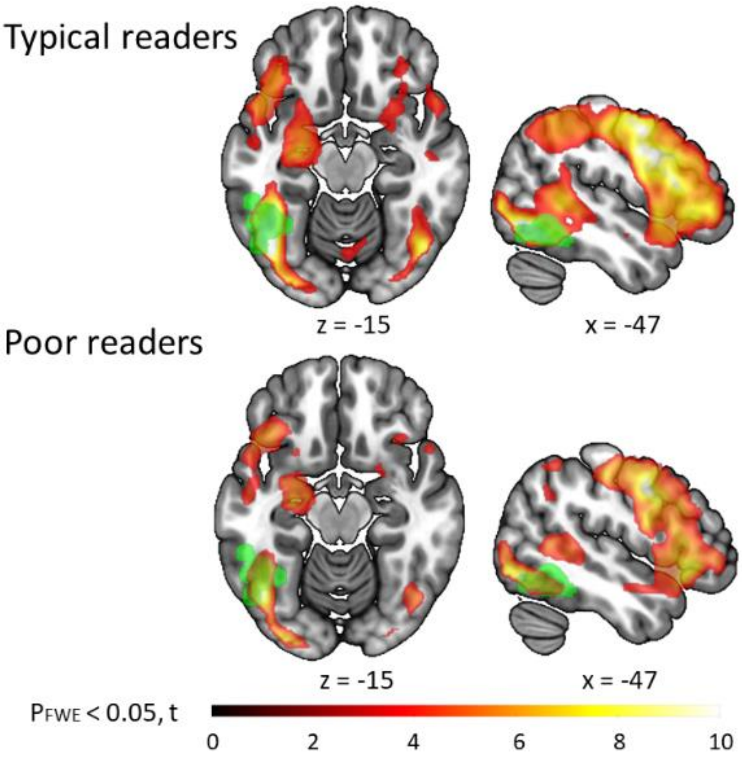


**Figure S4. VWFA** Localizer task results showed VWFA activation patterns for both typical (top) and poor readers (bottom). The VWFA mask is overlaid in green. $p_{CDT}$ = 0.001, $p_{\text{FWE}}$ corrected <0.05

#### Significant Clusters during the VWFA Localizer task in both Groups

**Table S4. Significant clusters for the contrast words vs checkerboards during the localizer task for typical readers**

| **Cluster** | | **Peak** | | **MNI coordinates** | | | **Location** |
|---|---|---|---|---|---|---|---|
| p(FDR-corr) | Cluster size | p(FWE-corr) | T | x {mm} | y {mm} | z {mm} | |
| **0** | **4011** | **0** | **14.547** | **-34** | **-90** | **-8** | |
| | | 0 | 12.126 | -44 | -66 | -18 | **Left inferior occipital gyrus** |
| | | 0 | 11.259 | -24 | -90 | -2 | Left occipital fusiform gyrus |
| **0** | **1891** | **0** | **11.115** | **36** | **-86** | **-8** | **Right inferior occipital gyrus** |
| | | 0 | 10.733 | 32 | -92 | -2 | Right inferior occipital gyrus |
| **0** | **4932** | **0.002** | **8.801** | **-42** | **8** | **26** | |
| | | 0.017 | 7.508 | -52 | 2 | 28 | **Left Inferior frontal gyrus** |
| | | 0.03 | 7.132 | -50 | 18 | 16 | |
| **0** | **1537** | **0.228** | **5.8** | **-50** | **-40** | **44** | |
| | | 0.28 | 5.655 | -28 | -58 | 48 | |
| | | 0.355 | 5.483 | -44 | -46 | 56 | Left superior parietal lobule |
| **0.002** | **347** | **0.275** | **5.67** | **54** | **-10** | **50** | |
| | | 0.415 | 5.362 | 38 | -12 | 64 | **Right postcentral gyrus** |
| | | 0.603 | 5.034 | 38 | -28 | 66 | Right precentral gyrus |
| **0.007** | **263** | **0.28** | **5.656** | **-30** | **-18** | **-6** | |
| | | 0.313 | 5.576 | -36 | -18 | -20 | **Left putamen** |
| | | 0.997 | 3.884 | -32 | -6 | -10 | |
| **0** | **480** | **0.371** | **5.448** | **-2** | **2** | **72** | |
| | | 0.733 | 4.819 | -8 | 0 | 60 | **Supplementary motor cortex** |
| | | 0.877 | 4.538 | -6 | 14 | 48 | |
| **0.011** | **231** | **0.487** | **5.23** | **50** | **-32** | **-2** | **Right middle temporal gyrus** |

Only clusters conforming to $p_{CDT} = 0.001$, $p_{\mathrm{FWEc}} < 0.05$, and $k \geq 231$ are listed.

**Table S5. Significant clusters for the contrast words vs checkerboards during the localizer task for poor readers**

| **Cluster** | | **Peak** | | **MNI coordinates** | | | **Location** |
|---|---|---|---|---|---|---|---|
| p(FWE-corr) | Cluster size | p(FWE-corr) | T | x {mm} | y{mm} | z {mm} | |
| **0** | **2224** | **0** | **12.94** | **-24** | **-98** | **-8** | |
| | | 0 | 12.48 | -40 | -80 | -8 | **Left inferior occipital gyrus** |
| | | 0 | 11.64 | -44 | -68 | -10 | |
| **0** | **14037** | **0** | **10.59** | **-48** | **0** | **38** | |
| | | 0.001 | 9.858 | -44 | 6 | 32 | **Left precentral gyrus** |
| | | 0.008 | 8.223 | -48 | 26 | -4 | |
| **0** | **3168** | **0.001** | **9.589** | **34** | **-90** | **-8** | |
| | | 0.004 | 8.623 | 30 | -94 | -2 | **Right inferior occipital gyrus** |
| | | 0.025 | 7.4 | 32 | -64 | -50 | |
| **0** | **796** | **0.02** | **7.537** | **-30** | **-58** | **56** | |
| | | 0.054 | 6.855 | -36 | -66 | 54 | **Left superior parietal lobule** |
| | | 0.548 | 5.159 | -44 | -52 | 54 | Left angular gyrus |
| **0.002** | **426** | **0.096** | **6.469** | **56** | **2** | **42** | **Right precentral gyrus** |
| | | 0.341 | 5.564 | 42 | -2 | 32 | |
| | | 0.953 | 4.309 | 34 | 6 | 36 | Right middle frontal gyrus |
| **0.016** | **282** | **0.861** | **4.586** | **46** | **8** | **-22** | |
| | | 0.912 | 4.456 | 48 | 24 | -6 | |
| | | 0.934 | 4.382 | 34 | 22 | -14 | Right inferior frontal gyrus<br>Posterior orbital gyrus |

Only clusters conforming to $p_{CDT} = 0.001$, $p_{\mathrm{FWEc}} < 0.05$, and $k \geq 282$ are listed.

### 2.2 No-feedback clusters during contrast regulation vs baseline pre and post runs

**Table S6**. **Significant activation clusters for the contrast regulation vs baseline during pre-training no-feedback runs for typical readers**

| **Cluster** | | **Peak** | | **MNI coordinates** | | | **Location** |
|---|---|---|---|---|---|---|---|
| p(FWE-corr) | Cluster size | p(FWE-corr) | T | x {mm} | y {mm} | z {mm} | |
| **0** | **1495** | **0.208** | **5.825** | **-40** | **16** | **22** | |
| | | 0.403 | 5.348 | -48 | 12 | 26 | **Left inferior frontal gyrus** |
| | | 0.491 | 5.185 | -54 | -6 | 38 | Left precentral gyrus |
| **0** | **599** | **0.214** | **5.805** | **-6** | **16** | **56** | |
| | | 0.568 | 5.054 | -4 | 6 | 62 | **Supplementary motor cortex** |
| | | 0.905 | 4.429 | -2 | 20 | 50 | |
| **0.003** | **384** | **0.38** | **5.393** | **20** | **-58** | **-24** | **Right cerebellum exterior** |
| | | 0.668 | 4.89 | 14 | -62 | -18 | |
| **0.039** | **218** | **0.587** | **5.022** | **24** | **-82** | **6** | |
| | | 0.957 | 4.249 | 12 | -96 | 8 | Right occipital pole |

Only clusters conforming to $p_{CDT} = 0.001$, $p_{\mathrm{FWEc}} < 0.05$, and $k \geq 218$ are listed.

**Table S7. Significant activation clusters for the contrast regulation vs baseline during pre-training no-feedback runs for poor readers**

| Cluster | | Peak | | MNI coordinates | | | Location |
|---|---|---|---|---|---|---|---|
| p(FWE-corr) | Cluster size | p(FWE-corr) | T | x {mm} | y {mm} | z {mm} | |
| **0.24** | **117** | **0.192** | **5.951** | **32** | **-98** | **2** | |
| | | 0.321 | 5.573 | 42 | -90 | -2 | **Right occipital pole** |
| | | 0.939 | 4.327 | 28 | -98 | 12 | Right inferior occipital gyrus |
| **0.00** | **408** | **0.240** | **5.789** | **-36** | **-94** | **6** | **Left inferior occipital gyrus** |
| | | 0.315 | 5.589 | -48 | -78 | -4 | |
| | | 0.512 | 5.186 | -14 | -104 | 4 | Left occipital pole |

Only clusters conforming to $p_{CDT}$ = 0.001, $p_{\mathrm{FWEc}}$ < 0.05, and k ≥ 408 are listed.

**Table S8. Significant activation clusters for the contrast regulation vs baseline during post-training no-feedback runs for typical readers**

| Cluster | | Peak | | MNI coordinates | | | Location |
|---|---|---|---|---|---|---|---|
| p(FWE-corr) | Cluster size | p(FWE-corr) | T | z {mm} | y {mm} | z {mm} | |
| **0** | **3273** | **0.006** | **8.474** | **-38** | **36** | **4** | |
| | | 0.031 | 7.253 | -58 | 18 | 24 | |
| | | 0.043 | 7.031 | -40 | 8 | 32 | Left middle frontal gyrus |
| **0.007** | **322** | **0.198** | **5.986** | **34** | **-70** | **-22** | **Right cerebellum exterior** |
| | | 0.491 | 5.281 | 22 | -60 | -24 | |
| | | 0.944 | 4.364 | 12 | -62 | -20 | |
| **0** | **671** | **0.278** | **5.74** | **-24** | **-18** | **0** | |
| | | 0.494 | 5.275 | -14 | -16 | 0 | |
| | | 0.658 | 4.99 | -16 | 6 | 2 | Left thalamus proper |
| **0.035** | **220** | **0.396** | **5.463** | **4** | **22** | **38** | |
| | | 0.625 | 5.046 | 12 | 24 | 36 | |
| | | 1 | 3.682 | -6 | 16 | 44 | **Supplementary motor cortex** |
| **0.038** | **215** | **0.505** | **5.254** | **-62** | **-30** | **2** | **Left superior temporal gyrus** |
| | | 0.777 | 4.778 | -54 | -30 | 0 | |
| | | 0.996 | 3.942 | -48 | -38 | 2 | |

Only clusters conforming to $p_{CDT}$ = 0.001, $p_{\mathrm{FWEc}}$ < 0.05, and k ≥ 215 are listed.

**Table S9. Significant activation clusters for the contrast regulation vs baseline during post-training no-feedback runs for poor readers**

| **Cluster** | | **Peak** | | **MNI coordinates** | | | **Location** |
|---|---|---|---|---|---|---|---|
| p(FWE-corr) | Cluster size | p(FWE-corr) | T | x {mm} | y {mm} | z {mm} | |
| **0** | **1298** | **0.019** | **7.678** | **12** | **18** | **40** | |
| | | 0.111 | 6.461 | -2 | 14 | 54 | Left Supplementary Motor Cortex |
| | | 0.145 | 6.275 | 2 | 18 | 44 | |
| **0** | **2707** | **0.094** | **6.575** | **-54** | **12** | **0** | **Left inferior frontal gyrus** |
| | | 0.095 | 6.568 | -18 | -2 | 16 | |
| | | 0.18 | 6.125 | -40 | 14 | 2 | Left frontal operculum |
| **0.006** | **312** | **0.203** | **6.039** | **30** | **-66** | **-26** | **Right cerebellum exterior** |
| | | 0.35 | 5.633 | 22 | -68 | -24 | |
| **0.033** | **210** | **0.304** | **5.74** | **42** | **18** | **-2** | **right anterior insula** |

Only clusters conforming to $p_{CDT} = 0.001$, $p_{\text{FWEc}} < 0.05$, and $k \geq 210$ are listed.

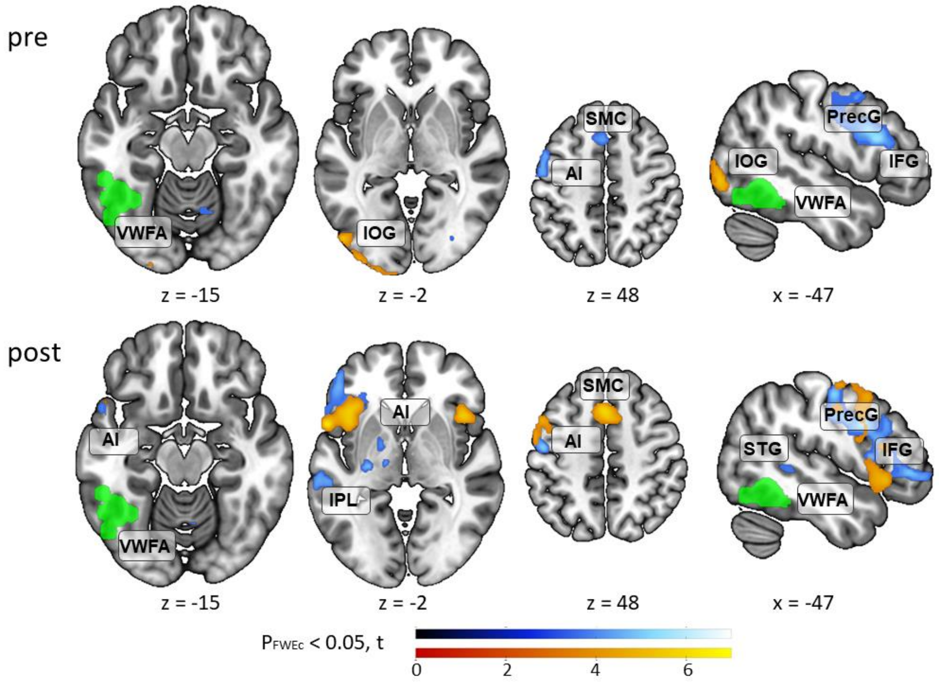


**Figure S5**. **Activations for the contrast regulation vs baseline during pre- and post-training no-feedback runs in typical and poor readers.** During pre-training no-feedback runs, significant activations were observed for participants with typical reading skills in the left inferior frontal gyrus (IFG), left precentral gyrus (PrecG), supplementary motor cortex (SMC), the right anterior insula (AI), and right occipital pole (cold colors), and for participants with poor reading skills in the bilateral occipital gyri (IOG) (warm colors). During post-training no-feedback runs, significant activations were observed for participants with typical reading skills in the left middle frontal gyrus (MFG), left superior temporal gyrus (STG), left inferior parietal lobe, left thalamus, the right supplementary motor cortex (SMC), and the left inferior frontal gyrus (IFG) (cold colors). For participants with poor reading skills in the left supplementary motor cortex (SMC), right anterior insula (AI), and left inferior frontal gyrus (IFG) (warm colors). Green: VWFA mask. Cluster defining threshold $p_{CDT} = 0.001$, $p_{\text{FWEc}} < 0.05$.

.

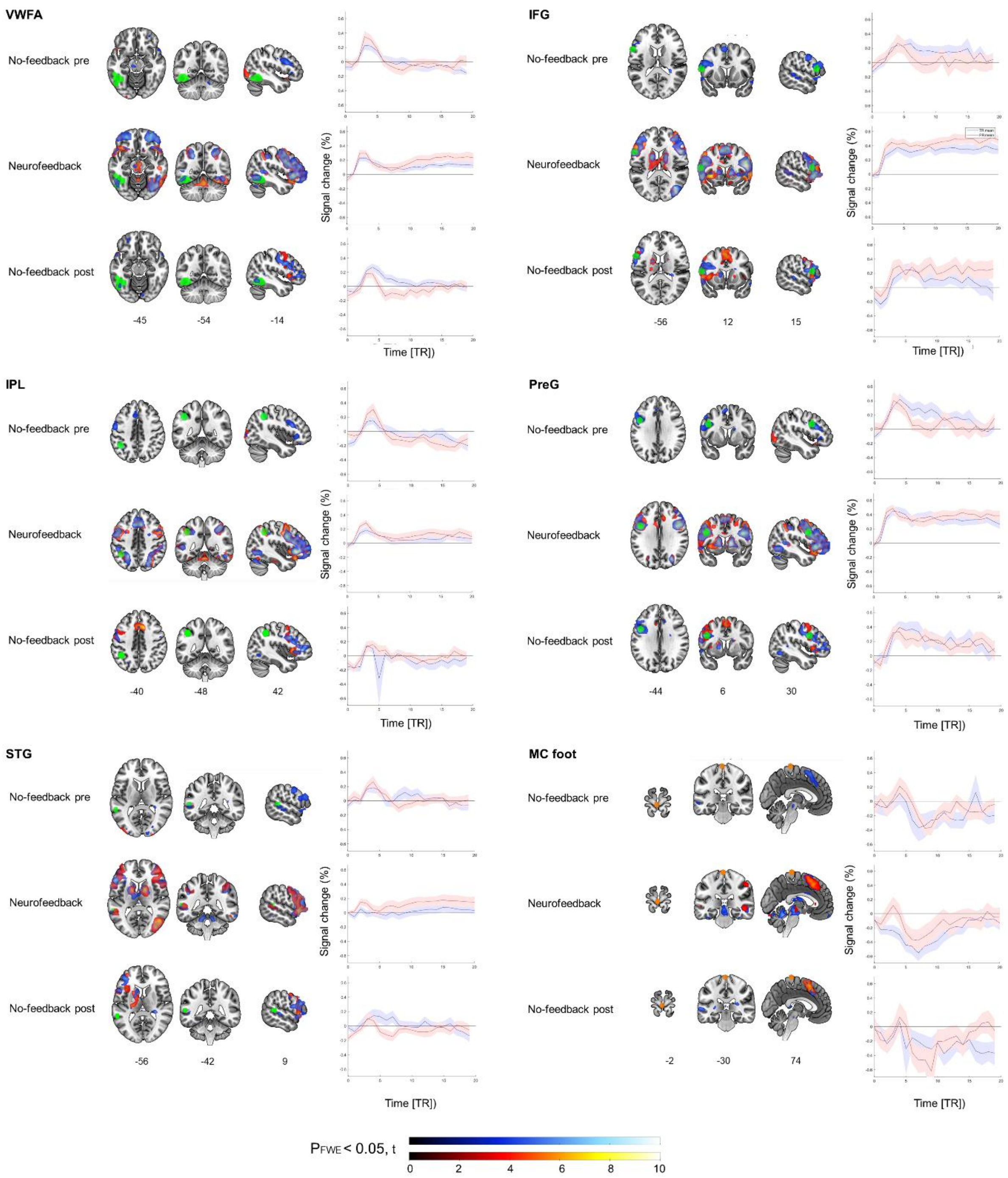


**Figure S6. Whole-brain activation as well as time courses in core regions of interest were analyzed during neurofeedback training and the pre- and post-training no-feedback runs.** These regions of interest related to reading (depicted in green) included the visual word form area (VWFA), inferior frontal gyrus (IFG), inferior parietal lobe (IPL), precentral gyrus (PreG), and superior temporal gyrus (STG). Additionally, we examined a region unrelated to reading, the foot area of the motor cortex (MC foot depicted in orange). For the region of interest, the brain slices on the left illustrate the activation clusters of participants with typical reading skills (blue) and poor reading skills (red). To the right of the brain images, the extracted percent signal change (PSC) time courses of each cluster are displayed for individuals with typical (blue) or poor (red) reading skills. Activity was averaged across all neurofeedback training runs.

#### 2.3 Additional Analyses with Percent Signal Change Time Courses

To examine group differences in sustained VWFA upregulation during the late phase, we compared the linear fits of VWFA percent signal change (PSC) across neurofeedback runs using ANCOVA. The dependent variable was PSC, with time within the second phase (20 seconds/10 TRs) and training timepoint (pre no-feedback, mean of neurofeedback runs, and post no-feedback as a within-subject factor and group (typical vs. poor readers) as a between-subject factor. This approach assessed whether the trajectory of sustained VWFA activation differed between groups during the late phase of neurofeedback upregulation over the three training timepoints.

This analysis revealed a main effect of training timepoint ($F(2, 1080)= 197.83$, $p < 0.001$), a main effect of group ($F(1, 1080)= 15.90$, $p < 0.001$) and the interaction effect ($F(2, 1080)= 16.70$, $p < 0.001$). Post-hoc analysis showed that groups differed in the neurofeedback ($t(1080) = 6.98$, $p < 0.0001$)), but not in the pre-training ($t(1080)= 0.089$, $p = 0.63$) and post-training ($t(1080) = -0.56$, $p = 0.58$) no-feedback runs. Moreover, sustained activity differed between neurofeedback training compared to pre-training ($t(1080) = 17.64$, $p < 0.0001$) and post-training ($t(1080) = 16.78$, $p < 0.0001$) no-feedback runs. No difference between pre- and post-training no-feedback runs was found ($t(1080) = 0.87$, $p = 0.66$).

#### 2.4 Mental Strategies Separated into Six Subgroups

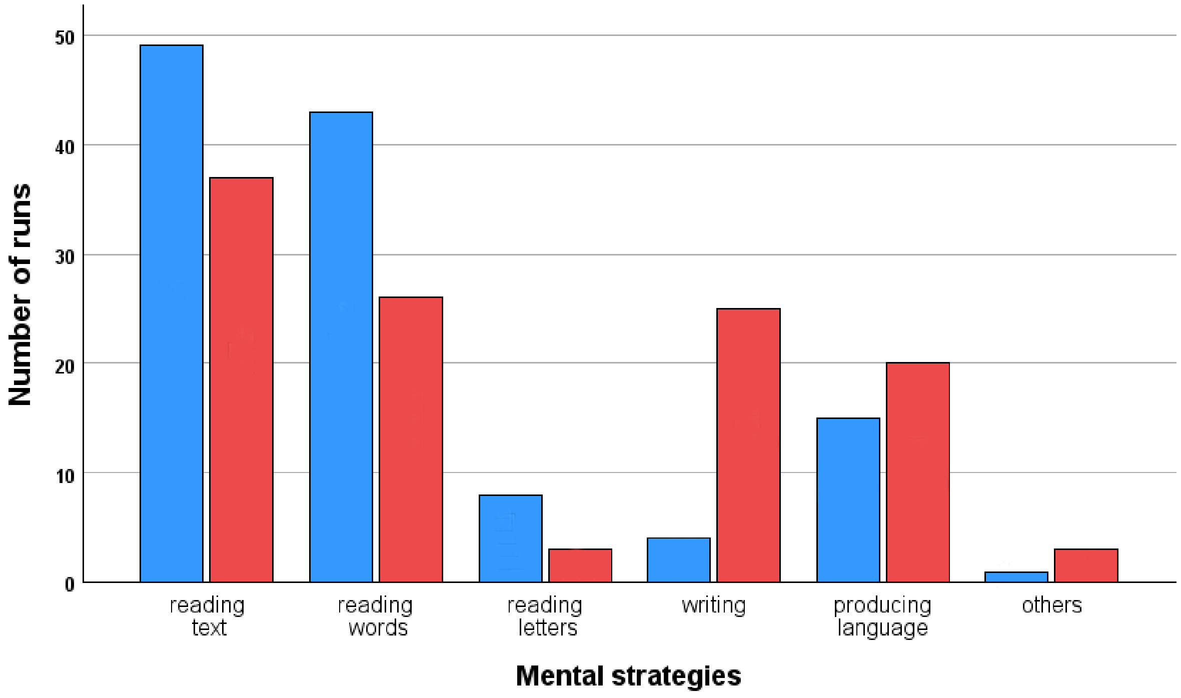


**Figure S7.** Strategies are grouped into six subcategories: reading text, reading words, reading letters, writing, producing language, and others for the group of participants with typical (blue) and poor reading skills (red). For the main analyses, these six subcategories were merged into two core strategies (see Figure 8).

#### 2.5 Comparison between Reading Fluency before and after Neurofeedback Training

For completeness, we also examined potential effects of time and/or neurofeedback training on reading skills by conducting a mixed ANOVA with the factors time (pre, post) and group (typical readers, TR; poor readers, PR) corrected for the covariate IQ. No significant interaction effect was found for pseudoword reading fluency ($F(1, 37) = 0.1$, $p = 0.753$). However, most measures showed significant improvement over time, and we observed a significant main effect of time for pseudoword reading fluency ($F(1, 37) = 21.01$, $p < 0.001$). No significant main effect of group was observed (see Figure S8).

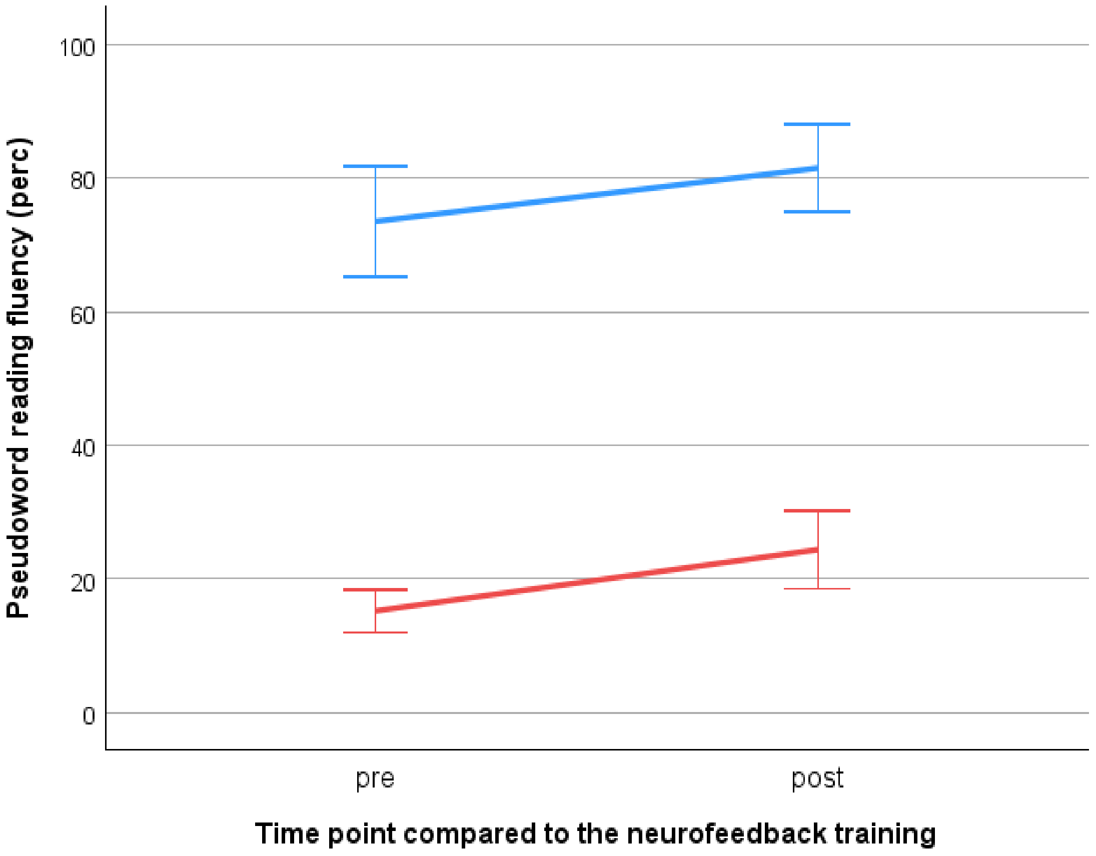


**Figure S8.** Reading fluency improved after neurofeedback training in both groups, i.e., participants with typical (blue) and poor reading skills (red). Error bars: +/- 1 SE.

As anticipated in this proof-of-concept study, our results reveal no significant difference in reading fluency improvements between adult participants with typical and poor reading skills; both groups showed substantial gains in their SLRT-II reading fluency scores for words and pseudowords. These improvements are most likely attributable to practice effects from repeated task exposure, as the same SLRT-II test version was administered twice in close succession. Consistent with this interpretation, our previous study (Haugg et al. 2023) also found no difference in reading score increases over time when comparing groups undergoing VWFA upregulation versus downregulation training
